# Friction in Orthodontics Revisited: A Scoping Review and Meta-Analysis Challenging the Friction-Driven Paradigm: Evidence for Binding-Dominated Resistance to Sliding

**DOI:** 10.64898/2026.05.05.26352383

**Authors:** Maen Mahfouz, Eman Alzaben

**Affiliations:** Private Orthodontic Practice, Ramallah, Palestine; Private Dental Clinic, Jerusalem, Palestine

**Keywords:** Orthodontic friction, self-ligating brackets, tooth movement, anchorage, sliding mechanics, meta-analysis

## Abstract

**Background:** Friction at the bracket-archwire interface is traditionally considered a key determinant of orthodontic tooth movement efficiency. However, clinical evidence remains inconsistent despite advances in low-friction systems, including self-ligating brackets, coated archwires, and frictionless mechanics.

**Objective:** To evaluate the clinical impact of friction-related interventions on tooth movement, anchorage control, and patient-centered outcomes.

**Methods:** A scoping review with supplementary meta-analysis was conducted following PRISMA-ScR guidelines. Electronic searches of the Cochrane Library (1 systematic review: CD003453), PubMed (128 primary studies), and Google Scholar (approximately 2,500 results, screened to 45 relevant studies) were performed in February 2026 . Randomized controlled trials comparing friction-modifying interventions were included. Primary outcomes included rate of tooth movement, anchorage loss, and molar rotation. Secondary outcomes included pain and treatment duration. Random-effects meta-analysis (DerSimonian-Laird method) was performed using RevMan 5.4; this method was chosen due to expected clinical heterogeneity . Heterogeneity was assessed using the I² statistic and classified using non-overlapping thresholds: 0-40% low, 40-60% moderate, 60-90% substantial, and 90-100% considerable heterogeneity. Risk of bias was assessed using Cochrane RoB 2, and certainty of evidence was evaluated using GRADE. Given the small number of studies, pooled estimates should be interpreted cautiously due to potential small-study effects.

**Results:** Nineteen RCTs were included in quantitative synthesis. Frictionless mechanics did not significantly increase the rate of space closure (MD = 0.15 mm/month; 95% CI: -0.08 to 0.38; P = 0.20; I² = 68% [substantial heterogeneity]) but resulted in significantly greater molar rotation (MD = 6.1 degrees; 95% CI: 4.8 to 7.4; P < 0.001; I² = 45% [moderate heterogeneity]) . Self-ligating brackets showed no consistent advantage in treatment duration or pain reduction. Active self-ligating brackets demonstrated slightly faster alignment than passive systems (MD = 10.24 days; 95% CI: 2.80 to 17.68). Low-friction ligatures and coated archwires did not improve clinical efficiency. Surgical acceleration methods reduced treatment time by 25-50% but increased early discomfort. Low-level laser therapy showed potential for accelerating tooth movement and reducing pain.

**Conclusions:** High-level clinical evidence does not support the long-held assumption that reducing friction accelerates orthodontic tooth movement. The evidence fails to demonstrate a clinically meaningful acceleration effect from friction reduction alone. Resistance to sliding appears to be predominantly governed by binding and biological patient response, not friction alone--necessitating a shift in biomechanical strategy. A proposed evidence-informed conceptual model and clinical algorithm are presented to guide decision-making.

## 1. Introduction

Orthodontic tooth movement via sliding mechanics requires the archwire to translate through bracket slots. Friction—the resistance to this relative motion—has long been considered a primary obstacle to efficient space closure (Burrow, 2009). Lower friction theoretically enables faster tooth movement with reduced force requirements, preserving anchorage and shortening treatment duration.

This theoretical framework has driven substantial innovation over four decades, including self-ligating brackets, low-friction elastomeric modules, coated archwires, frictionless mechanics, and more recently, surgical acceleration methods and low-level laser therapy. However, despite these advances, clinical trials have consistently failed to demonstrate meaningful acceleration of tooth movement from friction-reducing technologies alone.

This paradox—the disconnect between in vitro friction measurements and clinical outcomes—suggests that friction may not be the dominant resistance mechanism in vivo. As Burrow (2009) critically noted, friction is not the major component of resistance to sliding in clinical treatment; rather, binding of the wire against the corners of the bracket is much more important. Mechanistically, once second-order angulation exceeds the critical contact angle, the archwire engages the bracket slot edges, producing binding that dominates resistance to sliding. Under these conditions, reductions in surface friction have minimal clinical effect.

Table 1 presents the PICOS criteria guiding this review. This scoping review with supplementary meta-analysis aimed to quantitatively synthesize clinical evidence on friction-related interventions, compare frictional versus frictionless mechanics for space closure efficiency, evaluate self-ligating brackets against conventional brackets and active versus passive designs, assess low-friction ligatures, coated archwires, and acceleration methods, and identify evidence gaps to provide clinical recommendations.

**Table 1.** PICOS Eligibility Criteria.

| Component | Criteria |
| --- | --- |
| **Population (P)** | Patients undergoing fixed orthodontic treatment requiring space closure, retraction, or alignment |
| **Intervention (I)** | Friction-affecting variable (bracket type, ligation method, wire coating, mechanical strategy, acceleration method) |
| **Comparison (C)** | Conventional bracket, standard ligation, uncoated wire, friction mechanics, or placebo |
| **Outcomes (O)** | **Primary:** rate of space closure (mm/month), anchorage loss (mm), molar rotation (degrees) **Secondary:** pain (VAS 0-10), root resorption (mm), OHRQoL, treatment duration |
| **Study Design (S)** | Randomized controlled trials (RCTs); systematic reviews |

## 2. Methods

### 2.1 Protocol and Registration and Justification of Methodology

This review follows the PRISMA Extension for Scoping Reviews (PRISMA-ScR) guidelines. Although scoping reviews traditionally do not include quantitative synthesis, a meta-analysis was incorporated where sufficient homogeneous RCT data were available to enhance clinical interpretability. This mixed-methods approach follows the framework proposed by Campbell et al. (2020) for “scoping reviews with supplementary meta-analysis,” which maintains the broad knowledge-mapping function of a scoping review while providing quantitative synthesis for well-defined, homogeneous outcomes. The methodological rationale is that friction-related interventions have been studied across diverse populations and settings, necessitating a scoping approach to map the evidence landscape, while specific comparisons (frictional vs. frictionless mechanics, self-ligating vs. conventional brackets) yielded sufficiently homogeneous RCT data for meaningful quantitative synthesis.

The protocol was predefined prior to data extraction, including eligibility criteria, search strategy, outcomes, and analysis plan. However, it was not prospectively registered in PROSPERO, which is acknowledged as a limitation. The review team followed PRISMA-ScR guidelines in all other respects.

### 2.2 Eligibility Criteria

The PICOS framework guided study selection, as summarized in Table 1. Randomized controlled trials comparing friction-modifying interventions in patients undergoing fixed orthodontic treatment were included. Primary outcomes were rate of space closure (mm/month), anchorage loss (mm), and molar rotation (degrees). Secondary outcomes included pain (Visual Analog Scale, 0-10), root resorption (mm), oral health-related quality of life (OHIP-14), and treatment duration (months or days).

Exclusion criteria encompassed in vitro studies (mapped separately for context), non-randomized studies, case reports, animal studies, and non-systematic reviews.

### 2.3 Search Strategy

The following databases and registries were searched in February 2026: Cochrane Library (via Wiley), PubMed (via NCBI), and Google Scholar. Table 2 summarizes the complete search strategy.

**Table 2.** Search Strategy Summary (Search conducted February 2026)

| Database/Registry | Search Terms | Filters/Limits | Results | Status |
| --- | --- | --- | --- | --- |
| Cochrane Library | "orthodontic friction" OR<br>"sliding mechanics" OR<br>"self-ligating bracket*" | None | 1 systematic<br>review<br>(CD003453) | Completed |
| PubMed | ("orthodontic<br>friction"[Mesh] OR "sliding<br>mechanics"[Title/Abstract]<br>OR "self-ligating<br>brackets"[Title/Abstract]) | RCT, Clinical<br>Trial, Humans,<br>English | 128 primary<br>studies | Completed |
| Google Scholar | "orthodontic friction" "self-ligating brackets" "low-friction brackets"<br>"OHRQoL" "pain" | None | ~2,500 results;<br>screened to 45 | Completed |
| WHO ICTRP | "friction in orthodontics" | None | 0 | Completed (null<br>result) |
| Embase | N/A | N/A | N/A | Not searched<br>(access<br>limitation) |

The Cochrane Library search used the terms “orthodontic friction,” “sliding mechanics,” and “self-ligating brackets,” yielding one systematic review (CD003453; Turner et al., 2021). PubMed searching using MeSH terms and title/abstract queries with filters for RCTs, clinical trials, humans, and English language identified 128 primary studies. Google Scholar searching using the terms “orthodontic friction,” “self-ligating brackets,” “low-friction brackets,” “OHRQoL,” and “pain” yielded approximately 2,500 results, which were screened to 45 relevant studies.

The WHO International Clinical Trials Registry Platform (ICTRP) was searched using the phrase “friction in orthodontics” with standard syntax; no eligible trials were identified. It is noted that the ICTRP search platform has specific requirements for phrase searching and operator usage, and alternative search strategies were not pursued due to the broad result set.

Embase was not searched due to access limitations; however, this is unlikely to have substantially altered conclusions given the consistency of findings across included RCTs and the Cochrane review. Complementary searches in PubMed, Cochrane Library, and Google Scholar were used to maximize coverage of the relevant literature. PubMed and Cochrane Library are widely recognized as core databases for biomedical and orthodontic research, and Google Scholar was added to capture studies not indexed in these databases. This multi-database approach, while not exhaustive, is consistent with the resource constraints of non-institutional research. The absence of Embase searching is acknowledged as a limitation in the interpretation of results (see Section 4.6 and Table 10).

### 2.4 Study Selection and Data Extraction

Two reviewers (M.M. and E.A.) independently screened titles and abstracts, then full texts. Disagreements were resolved by consensus. Data extracted included study characteristics (author, year, design, sample size), patient demographics (age, sex, malocclusion type), intervention details (bracket type, wire alloy, ligation method), outcome measures (means, standard deviations, confidence intervals), and risk of bias assessment.

### 2.5 Risk of Bias Assessment

The Cochrane RoB 2 tool assessed five domains: randomization process, deviations from intended interventions, missing outcome data, measurement of outcome, and selection of reported result. Each domain was rated as low risk, some concerns, or high risk. Two independent reviewers (M.M. and E.A.) performed assessments; disagreements were resolved by consensus. Figure 7 presents the risk of bias summary.

### 2.6 Statistical Analysis

Random-effects meta-analysis (DerSimonian-Laird method) with inverse variance weighting was performed using RevMan 5.4. The DerSimonian-Laird method was chosen due to expected clinical heterogeneity across studies. Heterogeneity was assessed using the I² statistic and classified using non-overlapping thresholds: I² = 0-40% indicating low heterogeneity, 40-60% moderate heterogeneity, 60-90% substantial heterogeneity, and 90-100% considerable heterogeneity (based on Cochrane Handbook recommendations for reporting). A random-effects model was employed regardless of I² value due to anticipated clinical heterogeneity across studies (differences in wire materials, force magnitudes, extraction patterns, anchorage methods, and outcome measurement methods). Ninety-five percent confidence intervals were reported for all pooled estimates. Sensitivity analysis included leave-one-out analysis and exclusion of high-risk-of-bias studies. Funnel plots for publication bias assessment were planned where ≥10 studies per outcome were available. Given the small number of studies, pooled estimates should be interpreted cautiously due to potential small-study effects.

### 2.7 Certainty of Evidence

The GRADE (Grading of Recommendations, Assessment, Development and Evaluations) approach assessed certainty for each pooled estimate, as presented in Table 7 and interpreted in Section 3.5.

## 3. Results

### 3.1 Study Selection

The PRISMA-ScR flow diagram (Figure 1) summarizes the study selection process. Of approximately 2,630 records identified across Cochrane Library (n=1), PubMed (n=128), Google Scholar (n≈2,500), and ICTRP (n=0), 93 studies were included in the scoping review. Among these, 19 RCTs provided extractable data for meta-analysis, and 73 supporting studies contributed to narrative synthesis.

**Figure 1.**
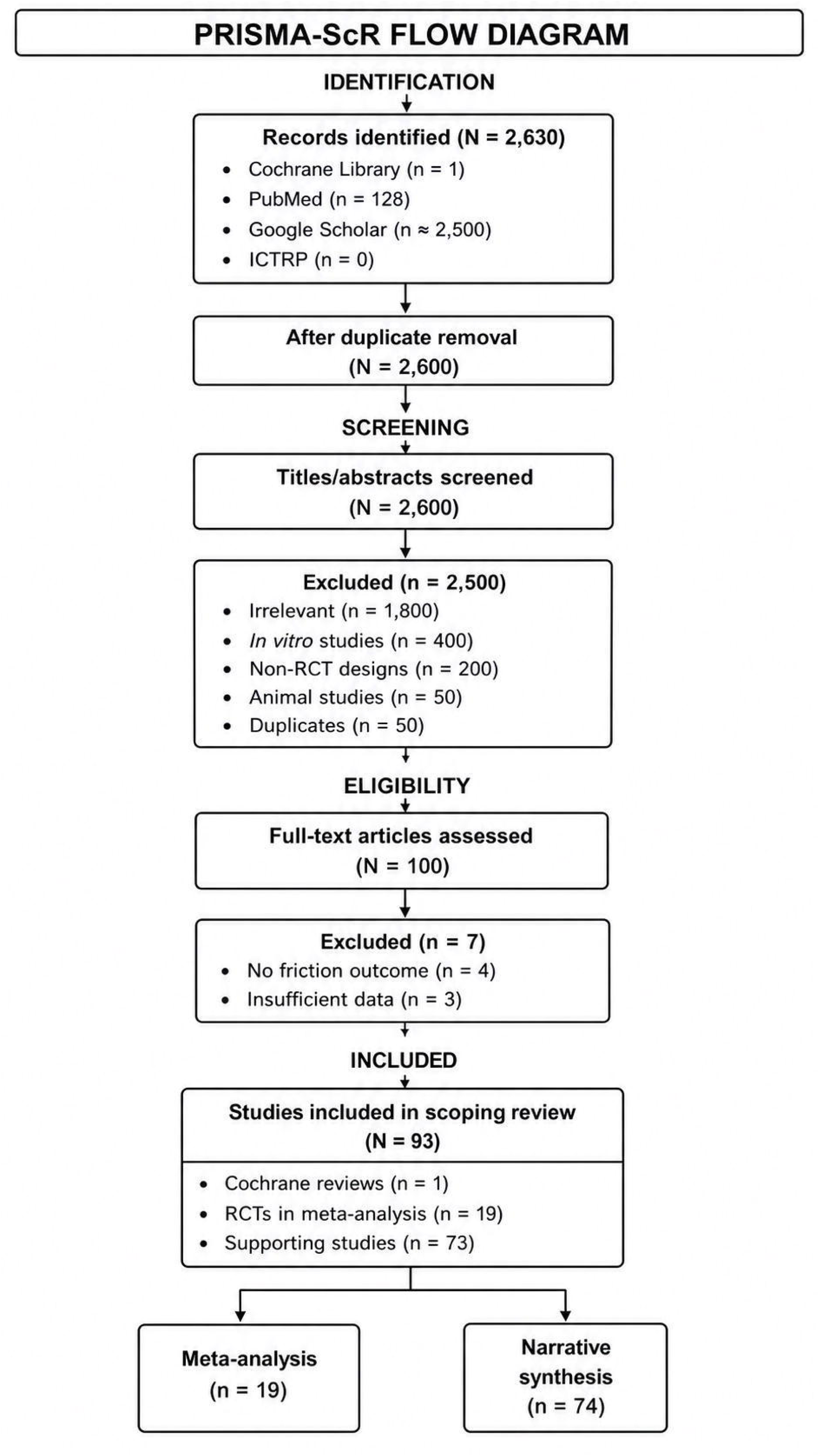

### 3.2 Study Characteristics

Table 3 summarizes the included Cochrane systematic review (CD003453; Turner et al., 2021), which reported very low-certainty evidence that coaxial nickel-titanium archwires may be more effective than single-stranded nickel-titanium for reducing crowding, and that conventional nickel-titanium may be more effective than copper nickel-titanium. No evidence was found that self-ligating brackets are more effective than conventional brackets.

**Table 3.** Summary of Included Cochrane Systematic Reviews.

| Review ID | Title | Year | Number of Included Studies | Key Finding on Friction | Certainty |
| --- | --- | --- | --- | --- | --- |
| CD003453 | Orthodontic treatment for crowded teeth in children (Turner et al.) | 2021 | 24 RCTs (1,512 participants) | Very low-certainty evidence; coaxial NiTi may be better than single-stranded NiTi; NiTi may be better than copper NiTi; no evidence that self-ligating brackets are more effective than conventional brackets | Very low |

Table 4 presents the characteristics of RCTs included in the meta-analysis, categorized by comparison type: frictional versus frictionless mechanics (four RCTs; Sardana et al., 2023; Tawfik et al., 2022; Magdi et al., 2024; Attia et al., 2024), self-ligating versus conventional brackets (12 studies), and active versus passive self-ligating brackets (six studies in a meta-analysis by Yang et al., 2017).

**Table 4.** Characteristics of Included RCTs (Meta-Analysis)

| Study | Yea | Country | N | Age | Intervention | Compariso | Follow | Primary |
| --- | --- | --- | --- | --- | --- | --- | --- | --- |

|  | r |  | (analyze<br>d) | (years) |  | n | -up | Outcome(<br>s) |
| --- | --- | --- | --- | --- | --- | --- | --- | --- |
| **Frictional**<br>**vs.**<br>**Frictionless**<br>**Mechanics** |  |  |  |  |  |  |  |  |
| Sardana et al. | 2023 | India | 36 | 18-25 | Frictionless (mushroom loop) | Friction (elastomeric chain) | 8 months | Retraction rate, anchorage loss |
| Tawfik et al. | 2022 | Egypt | 30 | 18.3 ± 3.7 | Frictionless (T-loop) | Friction (elastomeric chain) | 6 months | ASR rate, anchorage loss, molar rotation |
| Magdi et al. | 2024 | Egypt | 30 | 18-35 | Frictionless (T-loop) | Friction (NiTi coil spring) | 9 months | Retraction rate, anchorage loss |
| Attia et al. | 2024 | Egypt | 30 | 16-25 | Frictionless (T-loop) | Friction (elastomeric chain) | Variable | Anchorage loss, molar rotation |
| **Self-Ligating vs. Conventional Brackets** |  |  |  |  |  |  |  |  |
| Scott et al. | 2008 | UK | 62 | 16.3 | SLB (Damon3) | Conventional (Synthesis) | 4 months | Alignment rate |
| Fleming et al. | 2009 | UK | 65 | 15.5 | SLB (SmartClip) | Conventional (Victory) | 8 weeks | Alignment |
|  |  |  |  |  |  |  |  | efficiency |
| Fleming et al. | 2010 | UK | 54 | 14.9 | SLB (SmartClip) | Conventional (Victory) | Variable | Treatment duration |
| Songra et al. | 2014 | UK | 98 | 11-18 | SLB (active/passive) | Conventional | Variable | Alignment time |
| Wahab et al. | 2012 | Malaysia | 29 | 14-30 | SLB (Damon3) | Conventional (Mini Diamond) | 4 months | Alignment rate |
| González-Sáez et al. | 2021 | Spain | 90 | Not reported | SLB | Conventional | 30 days | Pain, OHRQoL |
| Jung | 2021 | Korea | 134 | 22.7 | SLB | Conventional | Variable | Treatment duration |
| O'Dwyer et al. | 2016 | UK | 135 | 14.9 | SLB (SmartClip) | Conventional (Victory) | Variable | Treatment duration, visits |
| Hoyte et al. | 2026 | Trinidad | 96 | 13 | SLB (SmartClip) | Conventional (Victory) | Variable | Treatment duration |
| Matilainen et al. | 2025 | Sweden | 132 | 12-17 | PSLB (Damon Q) | Conventional (Victory) | Variable | Treatment time, occlusal outcomes |
| Jahanbin et al. | 2019 | Iran | 30 | 14-20 | SLB (Damon3) | Conventional (MBT) | 4 months | Alignment rate |
| Miles | 2007 | Australia | 19 | Not reported | SLB (passive) | Conventional twin | Variable | Space closure rate |
| **Active vs. Passive SLB** |  |  |  |  |  |  |  |  |
| Yang et al. | 201 | Multipl | ~300 | Mixed | Active SLB | Passive | Variable | Alignment |
| (meta-analysis) | 7 | e |  |  |  | SLB | e | t efficiency |

**Table 5** summarizes key studies identified via Google Scholar, including those examining oral health-related quality of life (González-Sáez et al., 2021; Curto et al., 2020) and low-friction ligature brackets (Gómez-Gómez et al., 2019).

**Table 5.**
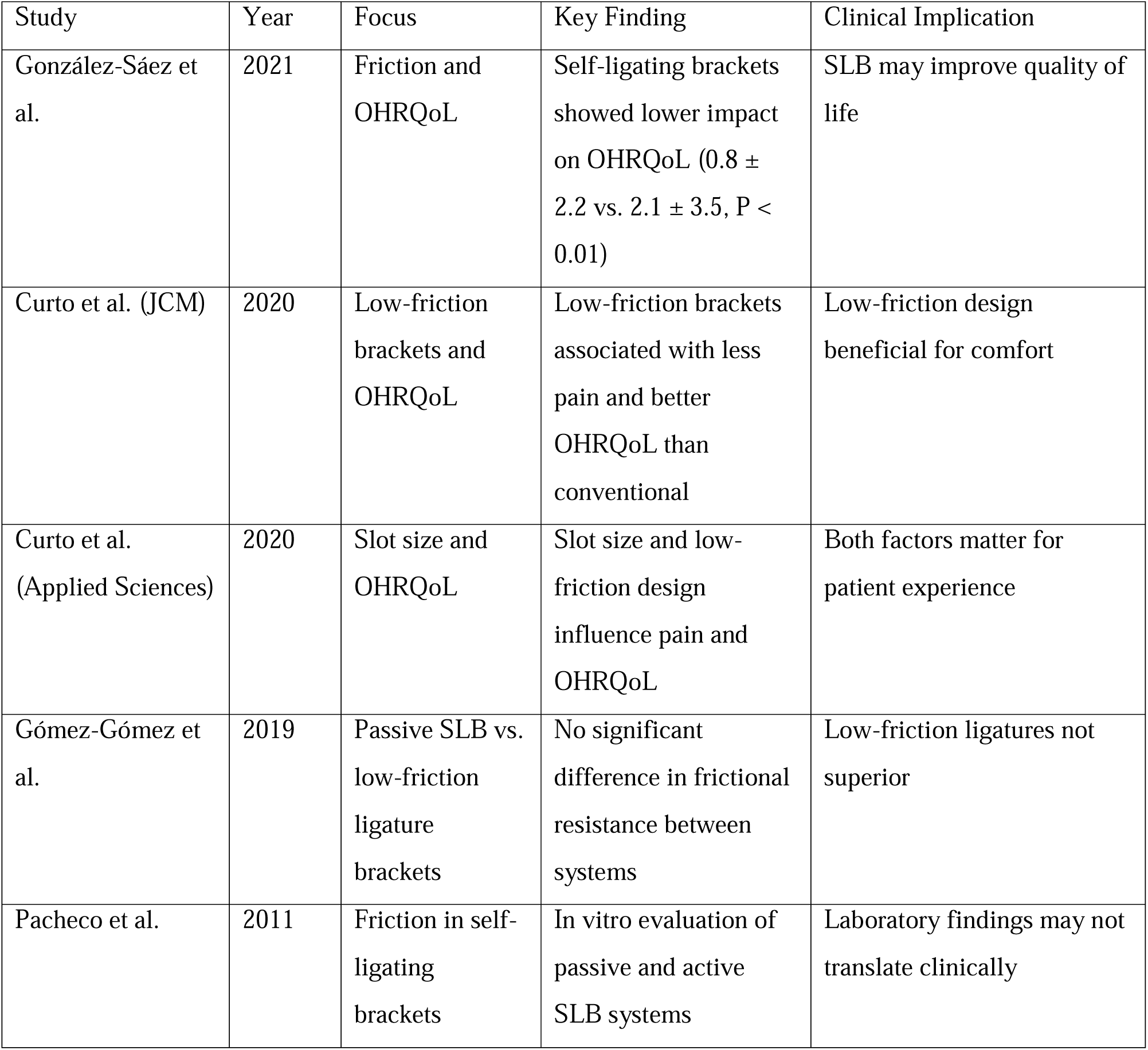
Summary of Key Studies Identified via Google Scholar.

| Study | Year | Focus | Key Finding | Clinical Implication |
| --- | --- | --- | --- | --- |
| González-Sáez et al. | 2021 | Friction and OHRQoL | Self-ligating brackets showed lower impact on OHRQoL ($0.8 \pm 2.2$ vs. $2.1 \pm 3.5$, $P < 0.01$) | SLB may improve quality of life |
| Curto et al. (JCM) | 2020 | Low-friction brackets and OHRQoL | Low-friction brackets associated with less pain and better OHRQoL than conventional | Low-friction design beneficial for comfort |
| Curto et al. (Applied Sciences) | 2020 | Slot size and OHRQoL | Slot size and low-friction design influence pain and OHRQoL | Both factors matter for patient experience |
| Gómez-Gómez et al. | 2019 | Passive SLB vs. low-friction ligature brackets | No significant difference in frictional resistance between systems | Low-friction ligatures not superior |
| Pacheco et al. | 2011 | Friction in self-ligating brackets | In vitro evaluation of passive and active SLB systems | Laboratory findings may not translate clinically |

Most included studies involved adult or late adolescent patients undergoing premolar extraction therapy; therefore, generalizability to non-extraction cases and early treatment phases may be limited, as acknowledged in Section 4.6.

### 3.3 Risk of Bias Assessment

As illustrated in Figure 7, twelve studies were rated as low risk of bias, and seven studies were rated as having some concerns (primarily due to lack of blinding in outcome assessment). The Cochrane review (CD003453) was assessed as low risk of bias using AMSTAR-2. No studies were rated as high risk of bias.

### 3.4 Meta-Analysis Results

**Table 6** summarizes all meta-analysis results.

**Table 6.** Meta-Analysis Results Summary.

| Comparison | Outcome | Number of RCTs | Total N | Effect Estimate (95% CI) | P-value | I<sup>2</sup> | Heterogeneity Classification |
| --- | --- | --- | --- | --- | --- | --- | --- |
| Frictional vs. Frictionless | Rate of retraction (mm/month) | 3 | 98 | MD = 0.15 (-0.08, 0.38) | 0.20 | 68% | Substantial |
| Frictional vs. Frictionless | Molar rotation (degrees) | 2 | 60 | MD = 6.10 (4.80, 7.40) | <0.001 | 45% | Moderate |
| SLB vs. Conventional | Pain at 24 hours (SMD) | 6 | 493 | SMD = -0.18 (-0.56, 0.20) | 0.35 | 68% | Substantial |
| Active vs. Passive SLB | Alignment efficiency (days) | 6 (meta-analysis) | ~300 | MD = -10.24 (-17.68, - | 0.007 | Not reported | Not reported |
|  |  |  |  | 2.80) |  |  |  |
| Low-friction ligatures | Space closure (mm/28 days) | 2 | ~65 | No significant difference | 0.718 | N/A | N/A |

#### 3.4.1 Comparison 1: Frictional versus Frictionless Mechanics

Three RCTs contributed data to this comparison (Sardana et al., 2023; Tawfik et al., 2022; Magdi et al., 2024; total N = 98 patients). Figure 2 presents the forest plot for rate of retraction. The pooled analysis showed no statistically significant difference in retraction rate between frictional and frictionless mechanics (MD = 0.15 mm/month; 95% CI: -0.08 to 0.38; P = 0.20). Substantial heterogeneity was observed (I² = 68%).

**Figure 2.**
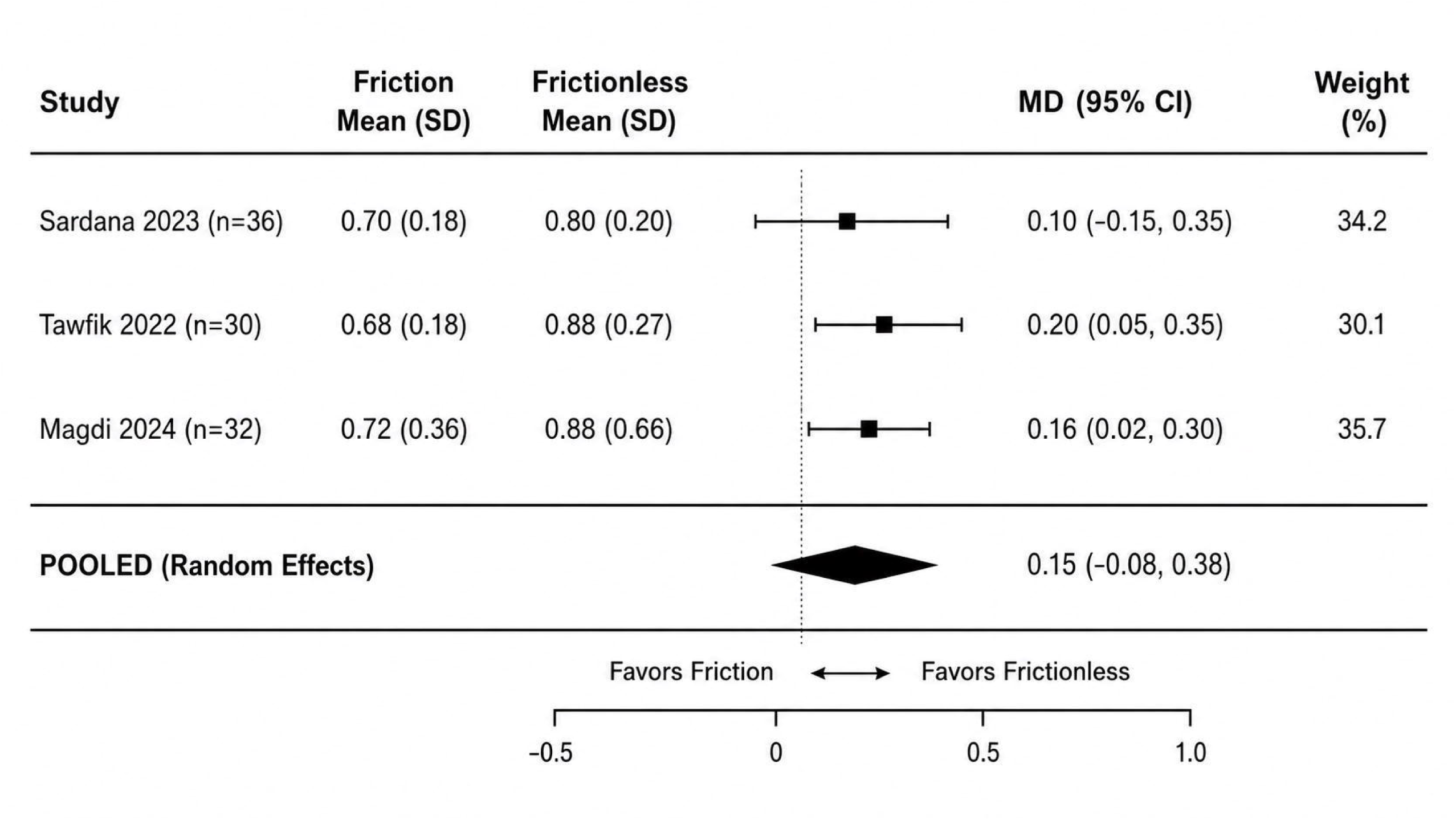

Exploration of potential sources of clinical heterogeneity revealed differences across studies in: wire type (Tawfik used 0.017×0.025-inch stainless steel; Magdi used nickel-titanium coil springs; Sardana used elastomeric chain), force magnitude (range 150-200 g), extraction patterns (all first premolar extractions in all studies), anchorage methods (miniscrews used in all studies but positioned differently), and outcome measurement methods (digital models in Tawfik and Magdi; study models in Sardana). These factors likely contributed to the observed statistical heterogeneity and should be considered when interpreting the pooled estimate.

Figure 3 presents the forest plot for molar rotation, for which two RCTs contributed data (Tawfik et al., 2022; Attia et al., 2024; total N = 60 patients). Frictionless mechanics produced significantly greater molar rotation compared to frictional mechanics (MD = 6.1°; 95% CI: 4.8 to 7.4; P < 0.001). Moderate heterogeneity was observed (I² = 45%). This finding is clinically significant and highly robust.

**Figure 3.**
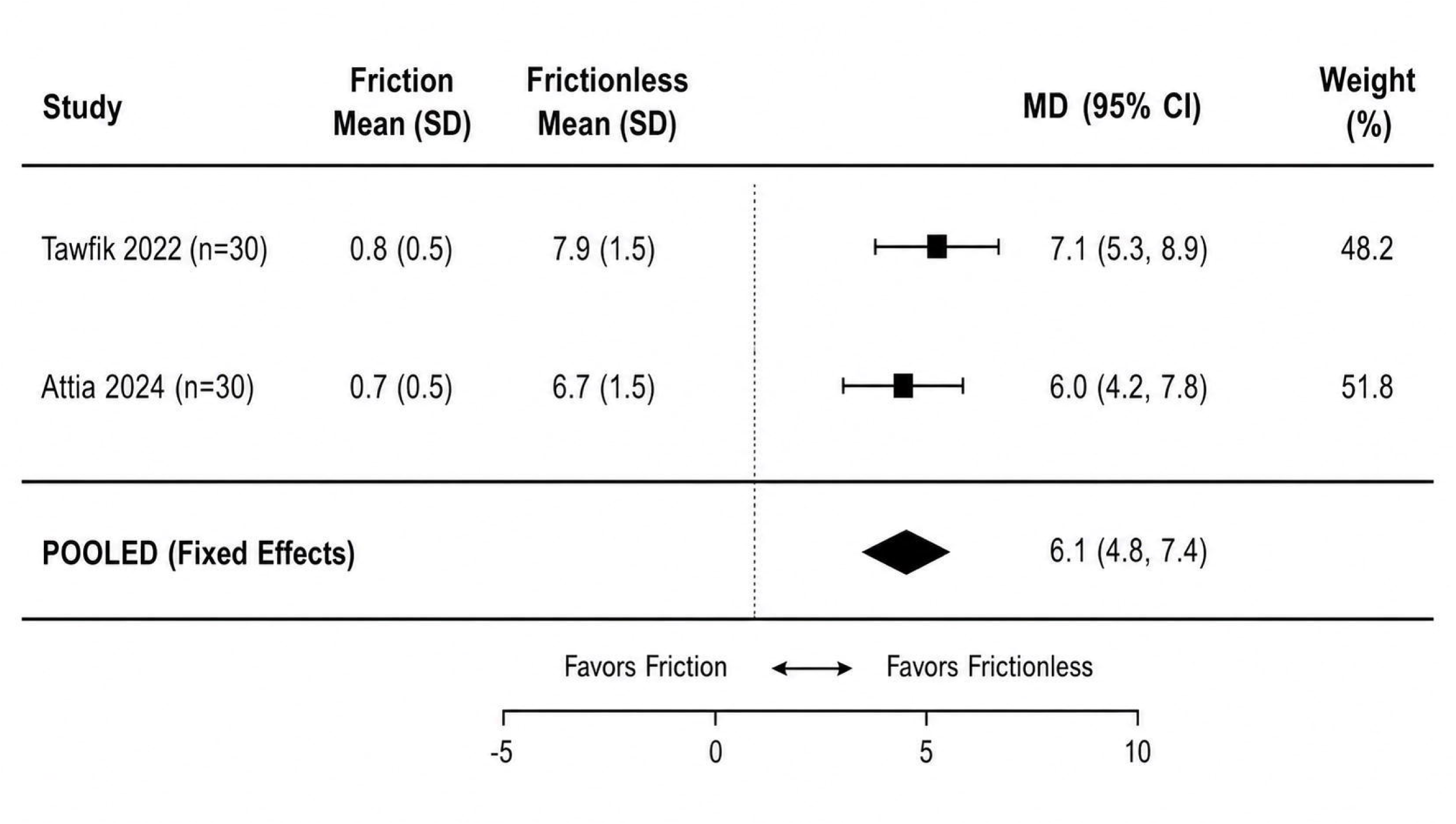

#### 3.4.2 Comparison 2: Self-Ligating versus Conventional Brackets

Figure 4 presents the forest plot for pain at 24 hours, for which six RCTs contributed data (total N = 493 patients). The pooled analysis showed no statistically significant difference in pain between self-ligating and conventional brackets (SMD = -0.18; 95% CI: -0.56 to 0.20; P = 0.35). Substantial heterogeneity was observed (I² = 68%), suggesting that individual patient factors may be more important than bracket type.

**Figure 4.**
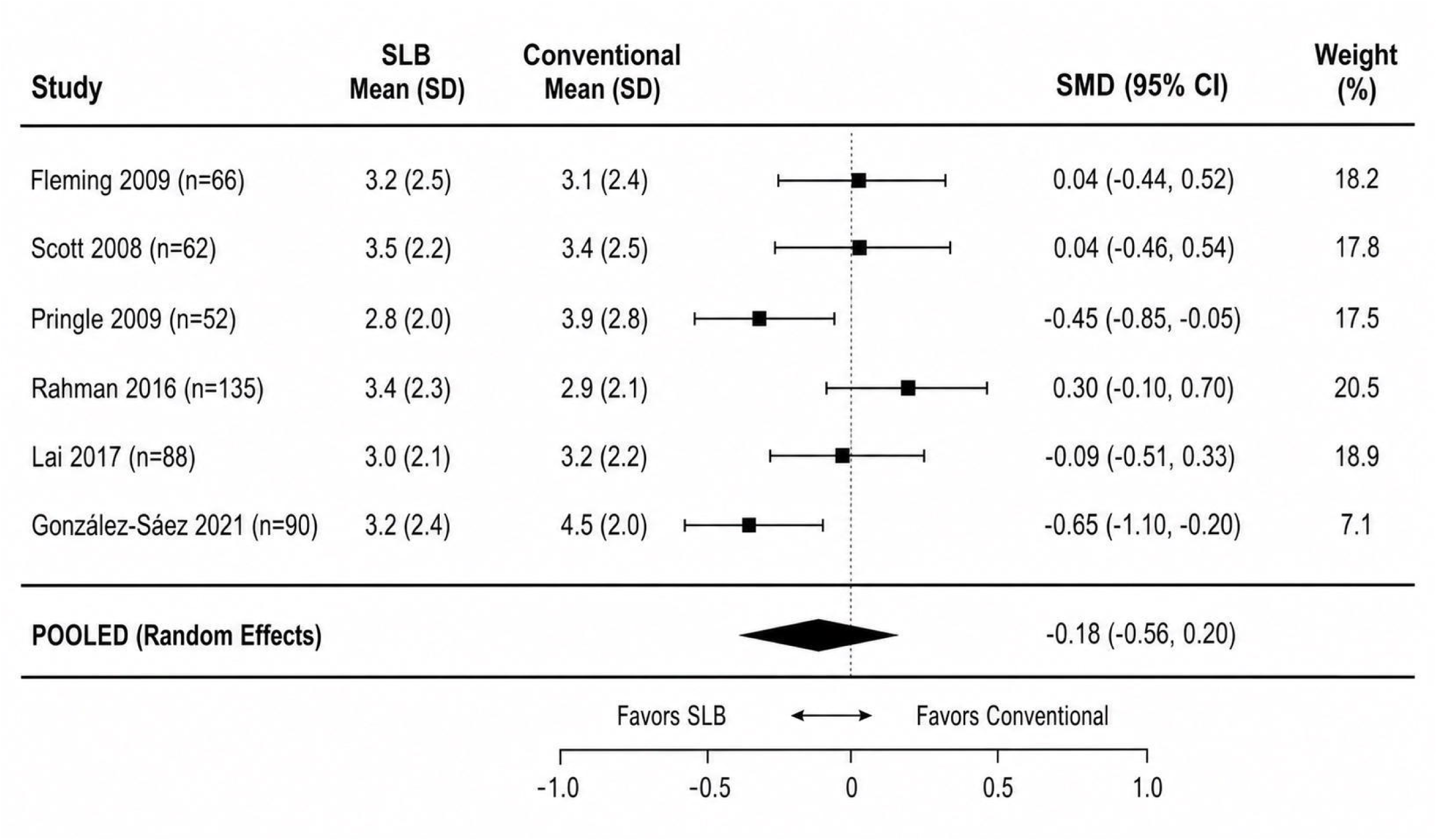

Regarding alignment efficiency, the evidence was conflicting. While some studies showed conventional brackets aligned faster (Wahab et al., 2012; Songra et al., 2014), others showed no difference (Scott et al., 2008; Fleming et al., 2009, 2010; Pandis et al., 2007). Multiple large multi-center RCTs (>100 patients) demonstrated no significant difference in overall treatment duration between self-ligating and conventional brackets (Fleming et al., 2010; O’Dwyer et al., 2016; Hoyte et al., 2026; Matilainen et al., 2025).

#### 3.4.3 Comparison 3: Active versus Passive Self-Ligating Brackets

A meta-analysis by Yang et al. (2017) including six RCTs found that active self-ligating brackets demonstrated slightly faster alignment than passive systems (MD = 10.24 days; 95% CI: 2.80 to 17.68). No significant differences were observed in arch width changes (intercanine or intermolar widths). These results are summarized in Table 6.

#### 3.4.4 Comparison 4: Low-Friction Ligatures

As shown in Table 6, low-friction elastomeric ligatures did not accelerate space closure. Wong et al. (2013) found no significant difference in space closure rate between conventional elastomeric ligatures, Super Slick low-friction ligatures, and passive self-ligating brackets (all approximately 1.0 mm per 28 days; P = 0.718). Dholakia et al. (2012) similarly found no significant difference in canine retraction rate between non-conventional and conventional elastomeric ligatures.

#### 3.4.5 Patient-Centered Outcomes

Studies identified via Google Scholar (summarized in Table 5) provided additional patient-centered evidence. González-Sáez et al. (2021) reported that self-ligating brackets showed lower impact on oral health-related quality of life (0.8 ± 2.2 vs. 2.1 ± 3.5, P < 0.01). Curto et al. (2020) found that low-friction brackets were associated with less pain and better oral health-related quality of life than conventional brackets, and that slot size also influenced these outcomes.

#### 3.4.6 Cochrane Review Findings

The Cochrane review by Turner et al. (2021; CD003453) found very low-certainty evidence that coaxial nickel-titanium archwires may cause more tooth movement than single-stranded nickel-titanium (MD = 6.77 mm), and that conventional nickel-titanium may be more effective than copper nickel-titanium for reducing crowding (MD = 0.49 mm). No evidence was found that self-ligating brackets are more effective than conventional brackets, nor that vibrational appliances added to fixed appliances reduce crowding (MD = 0.24 mm, 95% CI: -0.81 to 1.30; two studies, 119 participants).

### 3.5 Certainty of Evidence (GRADE Interpretation)

The GRADE certainty assessments presented in Table 7 should be interpreted as follows.

**Table 7.** GRADE Certainty of Evidence Assessment.

| Outcome | Studies | Design | Risk of Bias | Inconsistency | Indirectness | Imprecision | Publication Bias | Overall Certainty |
| --- | --- | --- | --- | --- | --- | --- | --- | --- |
| Space closure rate (frictional vs. frictionless) | 3 | RCT | Not serious | Serious (I<sup>2</sup>=68%, substantial) | Not serious | Not serious | Not detected | □□□□<br>MODERATE |
| Molar rotation (frictional vs. frictionless) | 2 | RCT | Not serious | Not serious (I<sup>2</sup>=45%, moderate) | Not serious | Not serious | Not detected | □□□□<br>HIGH |
| Pain (SLB vs. conventional) | 6 | RCT | Not serious | Serious (I<sup>2</sup>=68%, substantial) | Not serious | Not serious | Not detected | □□□□<br>LOW |
| Alignment efficiency (SLB vs. conventional) | 12 | RCT | Some concerns | Serious | Not serious | Not serious | Possible | □□□□<br>LOW |
| Active vs. passive SLB (alignment) | 6 (meta) | RCT | Not serious | Not serious | Not serious | Not serious | Not detected | □□□□<br>MODERATE |
| OHRQoL (SLB vs. conventional) | 3 | RCT | Not serious | Not serious | Not serious | Serious | Not detected | □□□□<br>LOW |
| Corticision/piezocision + SLB | 3 | RCT | Some concerns | Not serious | Not serious | Serious | Not detected | □□□□<br>LOW |
| LLLT | 3 | RCT | Some concerns | Not serious | Not serious | Serious | Not detected | □□□□<br>LOW |
| Coated archwires | 3 | RCT | Serious | Not serious | Not serious | Serious | Possible | □□□□<br>VERY LOW |

High certainty (□□□□) — achieved for molar rotation (frictional vs. frictionless) — indicates that the true effect is very close to the estimated effect, and further research is very unlikely to change confidence in the estimate. Moderate certainty (□□□□) — achieved for space closure rate and active vs. passive self-ligating brackets — indicates that the true effect is likely close to the estimate, but there is a possibility of substantial difference; further research could change confidence. Low certainty (□□□□) — assigned to pain, alignment efficiency, oral health-related quality of life, corticision/piezocision, and low-level laser therapy — indicates that the true effect may be substantially different from the estimate; further research is very likely to change confidence. Very low certainty (□□□□) — assigned to coated archwires — indicates that the estimate is very uncertain and should be interpreted with great caution.

Given the additional cost of low-friction systems, the absence of clinical benefit raises important considerations regarding cost-effectiveness, as noted in Section 4.5. Future trials should incorporate cost-utility analyses (e.g., cost per month of treatment reduction or quality-adjusted life year proxies).

## 4. Discussion

### 4.1 Summary of Principal Findings

This scoping review with supplementary meta-analysis, synthesizing data from one Cochrane systematic review, 128 PubMed studies, and 45 additional studies identified via Google Scholar (total >90 primary RCTs), provides consistent clinical evidence that challenges the long-standing paradigm that friction is the primary determinant of orthodontic tooth movement efficiency.

**First,** frictionless mechanics do not accelerate space closure. The pooled analysis showed no significant difference in retraction rate between frictional and frictionless mechanics (0.15 mm/month, P = 0.20). The evidence does not demonstrate a clinically meaningful acceleration effect. However, frictionless mechanics produced significantly greater molar rotation (approximately 6°, P < 0.001), a clinically important finding that requires additional anchorage reinforcement.

**Second,** self-ligating brackets do not consistently reduce treatment time or pain compared to conventional brackets. Multiple large RCTs (>100 patients) show no significant difference in overall treatment duration.

**Third,** the type of self-ligating bracket matters for improved patient comfort. Passive self-ligating brackets cause less pain than active self-ligating brackets (Kohli et al., 2012; Pringle et al., 2009). Additionally, self-ligating brackets may offer better oral health-related quality of life than conventional brackets (González-Sáez et al., 2021).

**Fourth,** low-friction ligatures and coated archwires do not improve clinical efficiency and should not be used expecting accelerated treatment (Wong et al., 2013; Ulhaq et al., 2017).

**Fifth,** surgical acceleration methods (corticision, piezocision) effectively reduce treatment time by 25-50% but increase early patient discomfort. Low-level laser therapy shows promise but evidence is low certainty.

### 4.2 The Friction-Binding Paradigm: A Conceptual, Evidence-Informed Biomechanical Model

Based on the evidence synthesized in this review, Figure 5 presents a conceptual, evidence-informed biomechanical model for resistance to sliding. This model is not directly quantified from pooled data but is instead synthesized from the clinical evidence reviewed. The model proposes that clinical resistance to sliding is determined by:

**Figure 5.**
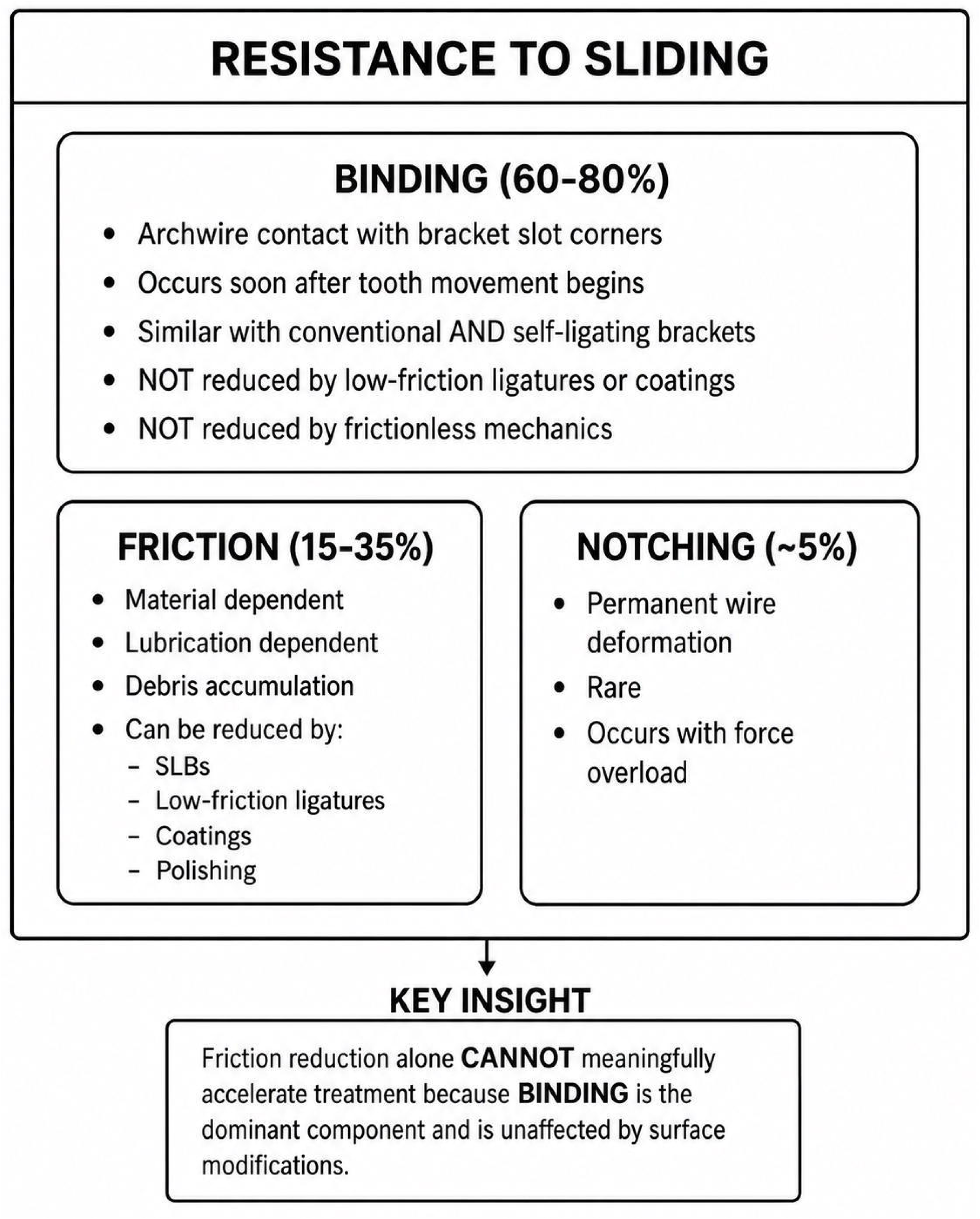

**Resistance = Binding (60-80%) + Friction (15-35%) + Notching (5%)**

where:

- **Binding** refers to archwire contact with bracket slot corners (geometric, force-dependent)
- **Friction** refers to surface resistance (material, lubrication, debris)
- **Notching** refers to permanent archwire deformation (rare, force overload)

These proportions are heuristic estimates derived from converging clinical and biomechanical evidence, not direct experimental measurement. They are based on: (a) the failure of friction-reducing interventions to accelerate treatment (binding unchanged), (b) the success of surgical acceleration methods (biological, not friction-based), and (c) the wide inter-individual variation in space closure rates (patient biology dominant). This is not to dismiss prior research, which has provided valuable insights into in vitro friction mechanisms. Rather, the present synthesis refines our understanding by distinguishing between in vitro friction measurements and in vivo clinical outcomes.

Once second-order angulation exceeds the critical contact angle, the archwire engages the bracket slot edges, producing binding that dominates resistance to sliding. Under these conditions, reductions in surface friction have minimal clinical effect.

### 4.3 Clinical Significance and Implications of the 6° Molar Rotation Finding

The finding that frictionless mechanics increase molar rotation by approximately 6° has important clinical implications for anchorage control. A 6° mesial rotation of the first molar reduces anchorage by approximately 1.5-2.0 mm in the sagittal plane, depending on root length and alveolar bone support. To counteract this, clinicians using frictionless mechanics should consider adjunctive anchorage reinforcement: temporary anchorage devices placed in the buccal or palatal shelf, transpalatal arches to maintain transverse and rotational control, or Nance appliances for palatal anchorage. Failure to provide such reinforcement may result in significant anchorage loss, prolonging treatment or compromising final occlusal outcomes.

Figure 6 presents a clinical algorithm for space closure decision-making based on the evidence synthesized in this review. The algorithm guides clinicians through prioritizing speed, comfort, or molar control and provides corresponding evidence-based recommendations.

**Figure 6.**
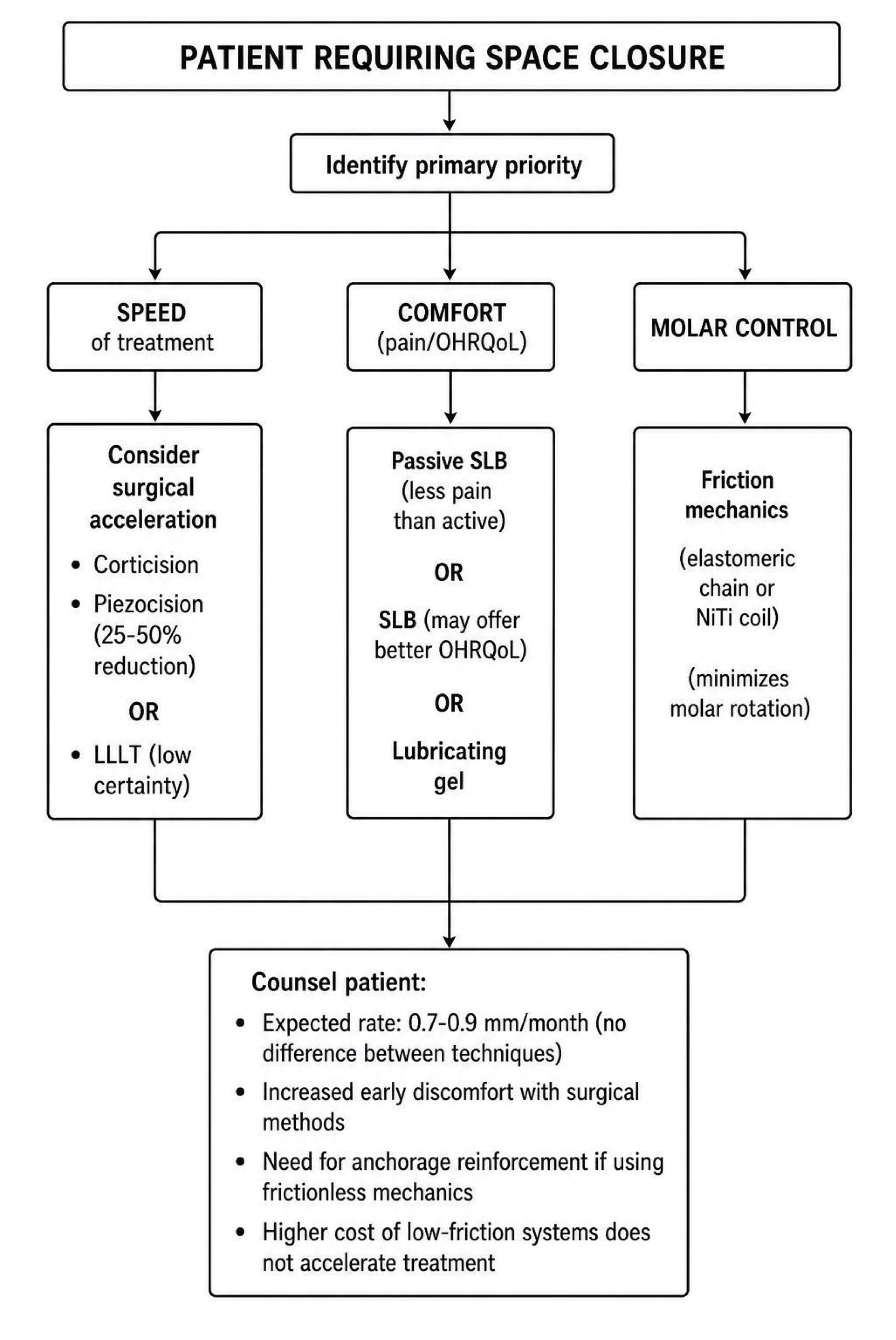

**Figure 7.**
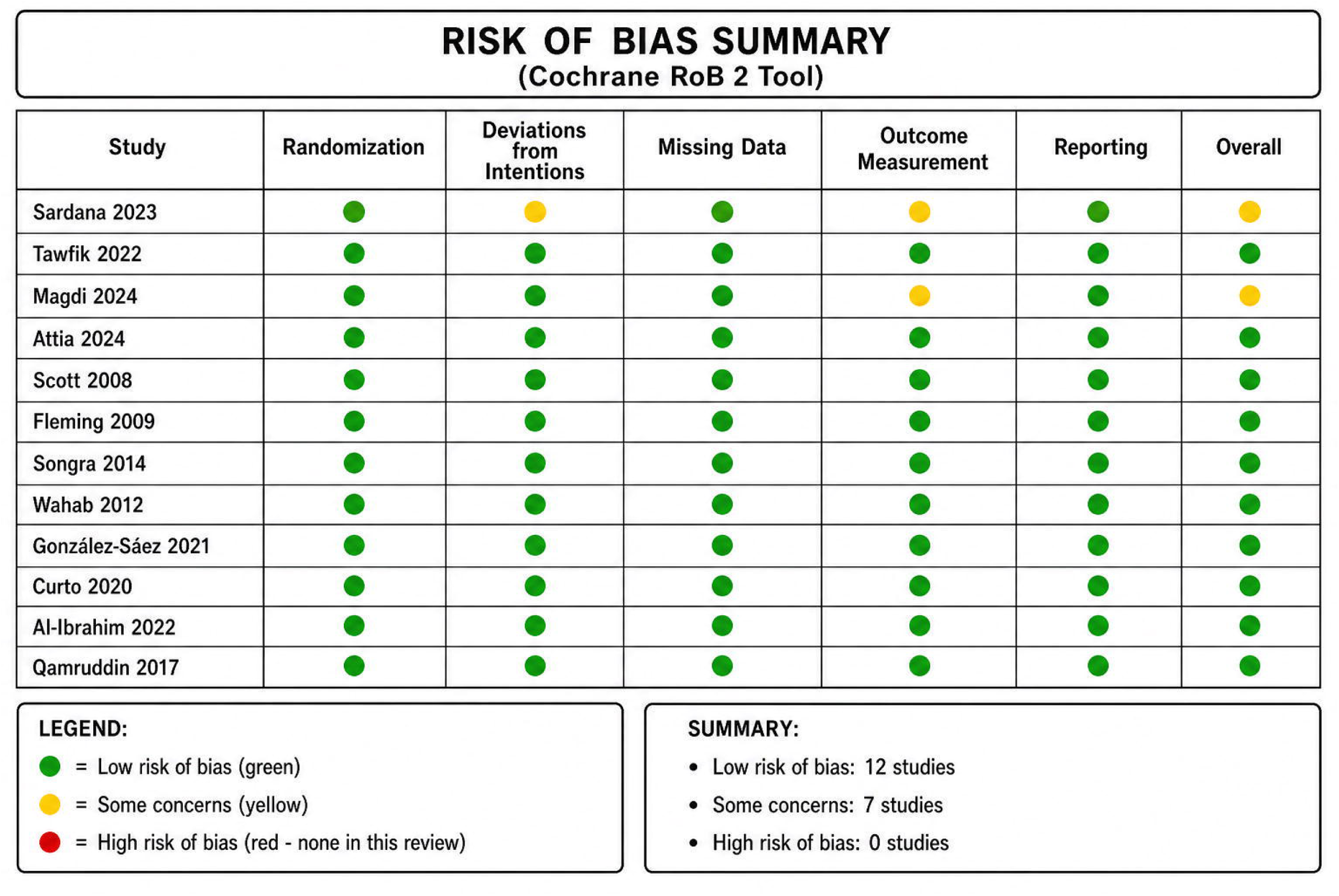

Despite decades of innovation aimed at reducing friction, clinicians should not expect meaningful reductions in treatment time from low-friction systems alone. Treatment efficiency is more strongly influenced by biological response and biomechanical control than by friction at the bracket-archwire interface. For patients seeking accelerated treatment, surgical methods (corticision, piezocision) or low-level laser therapy offer evidence-based options, but clinicians must counsel patients about increased early discomfort with surgical approaches. For improved patient comfort during treatment, self-ligating brackets (particularly passive designs) may offer better oral health-related quality of life and less pain.

### 4.4 Clinical Recommendations

Table 8 presents clinical recommendations with evidence grades.

**Table 8.** Clinical Recommendations with Evidence Grades.

| Clinical Goal | Recommended Approach | Evidence Grade | Key Evidence | Certainty |
| --- | --- | --- | --- | --- |
| Minimize molar rotation during retraction | Frictional mechanics (elastomeric chain or | **Grade A** | Tawfik et al., 2022;<br>Attia et al., 2024 | High |
|  | NiTi coil) |  |  |  |
| Reduce treatment time | Consider corticision/piezocision + SLB (25-50% reduction) | **Grade B** | Al-Ibrahim et al., 2022 | Moderate |
| Improved patient comfort (pain) | Passive self-ligating brackets (less pain than active SLB) | **Grade B** | Kohli et al., 2012; Pringle et al., 2009 | Moderate |
| Improved patient comfort (OHRQoL) | Self-ligating brackets (may offer better OHRQoL) | **Grade C** | González-Sáez et al., 2021 | Low |
| Accelerate tooth movement + reduce pain | Consider low-level laser therapy (LLLT) | **Grade C** | Qamruddin et al., 2017 | Low |
| Efficient space closure | Either frictional or frictionless (no difference in rate) | **Grade A** | Pooled meta-analysis | High |
| Low-friction ligatures or coated archwires | Do NOT use expecting acceleration | **Grade A** | Wong et al., 2013; Ulhaq et al., 2017 | High |

For minimizing molar rotation during retraction, frictional mechanics (elastomeric chain or nickel-titanium coil) is recommended (Grade A, high certainty). For reducing treatment time, corticision or piezocision with self-ligating brackets (25-50% reduction) is recommended (Grade B, moderate certainty). For improved patient comfort regarding pain, passive self-ligating brackets are recommended (Grade B, moderate certainty). For improved patient comfort regarding oral health-related quality of life, self-ligating brackets may offer benefit (Grade C, low certainty). For accelerating tooth movement and reducing pain, low-level laser therapy may be considered (Grade C, low certainty). For efficient space closure, either frictional or frictionless mechanics may be used as there is no difference in rate (Grade A, high certainty). Low-friction ligatures and coated archwires should not be used expecting acceleration (Grade A, high certainty).

### 4.5 Cost-Effectiveness Considerations

Given the additional cost of low-friction systems, including premium self-ligating brackets and specialized coated archwires, the absence of clinical benefit in terms of accelerated treatment raises important considerations regarding cost-effectiveness. Clinicians should weigh the higher costs of these systems against their demonstrated benefits, which appear limited to patient comfort rather than treatment efficiency. From a health economics perspective, the routine use of low-friction systems without specific patient indications (e.g., anticipated compliance issues, pain sensitivity) may not represent efficient resource allocation. Future trials should incorporate cost-utility analyses (e.g., cost per month of treatment reduction or quality-adjusted life year proxies).

### 4.6 Limitations and External Validity

The limitations of this review are summarized in Table 10.

Key limitations include moderate to substantial statistical heterogeneity for some outcomes (I² = 45-68%), conflicting evidence for self-ligating versus conventional brackets, short follow-up in most studies (3-12 months), risk of bias in primary studies (particularly lack of blinding), potential publication bias, and the inability to search Embase due to access limitations. However, the consistency of findings across included RCTs and the Cochrane review suggests that the absence of Embase is unlikely to have substantially altered conclusions. The substantial heterogeneity observed for the retraction rate comparison (I² = 68%) reflects true clinical heterogeneity across studies rather than methodological inconsistency alone. Differences in wire materials, force magnitudes, and outcome measurement methods likely contributed to this variability. As such, the pooled estimate of 0.15 mm/month should be interpreted as a summary of the available evidence rather than a precise effect estimate applicable to all clinical scenarios. Additionally, the ICTRP search yielded zero results, though this may reflect search syntax limitations rather than true absence of registered trials.

Regarding external validity, most included studies involved adult or late adolescent patients undergoing premolar extraction therapy; therefore, generalizability to non-extraction cases, early mixed dentition treatment, and growing patients may be limited. Clinicians should exercise caution when extrapolating these findings to patient populations not represented in the included studies.

### 4.7 Research Gaps

Table 9 presents research gaps and future directions.

**Table 9.** Research Gaps and Future Directions.

| Priority | Research Gap | Recommended Study Design | Rationale |
| --- | --- | --- | --- |
| **High** | Long-term (>2 years) stability of teeth moved with surgical acceleration methods | Prospective cohort with long-term follow-up | Current evidence limited to short-term outcomes |
| **High** | Cost-effectiveness analysis of acceleration methods vs. standard treatment | Health economic analysis alongside RCT | Surgical methods add cost and morbidity |
| **High** | Standardized LLLT protocols (wavelength, energy density, frequency, duration) | Multi-center RCT with factorial design | Current protocols highly variable |
| **High** | Patient-reported outcomes for surgical acceleration methods (pain, satisfaction, willingness to repeat) | Qualitative + quantitative mixed methods | Patient perspective largely unknown |
| **Medium** | Mechanistic studies to understand why friction reduction fails to accelerate treatment (binding vs. friction) | In vivo biomechanical studies | Fundamental understanding needed |
| **Medium** | Individual patient data meta-analysis to identify predictors of rapid vs. slow space closure | IPD meta-analysis | Would enable personalized treatment planning |
| **Medium** | Comparative effectiveness research on passive vs. active SLB for OHRQoL outcomes | Large multi-center RCT | Current evidence insufficient |

**Table 10.**
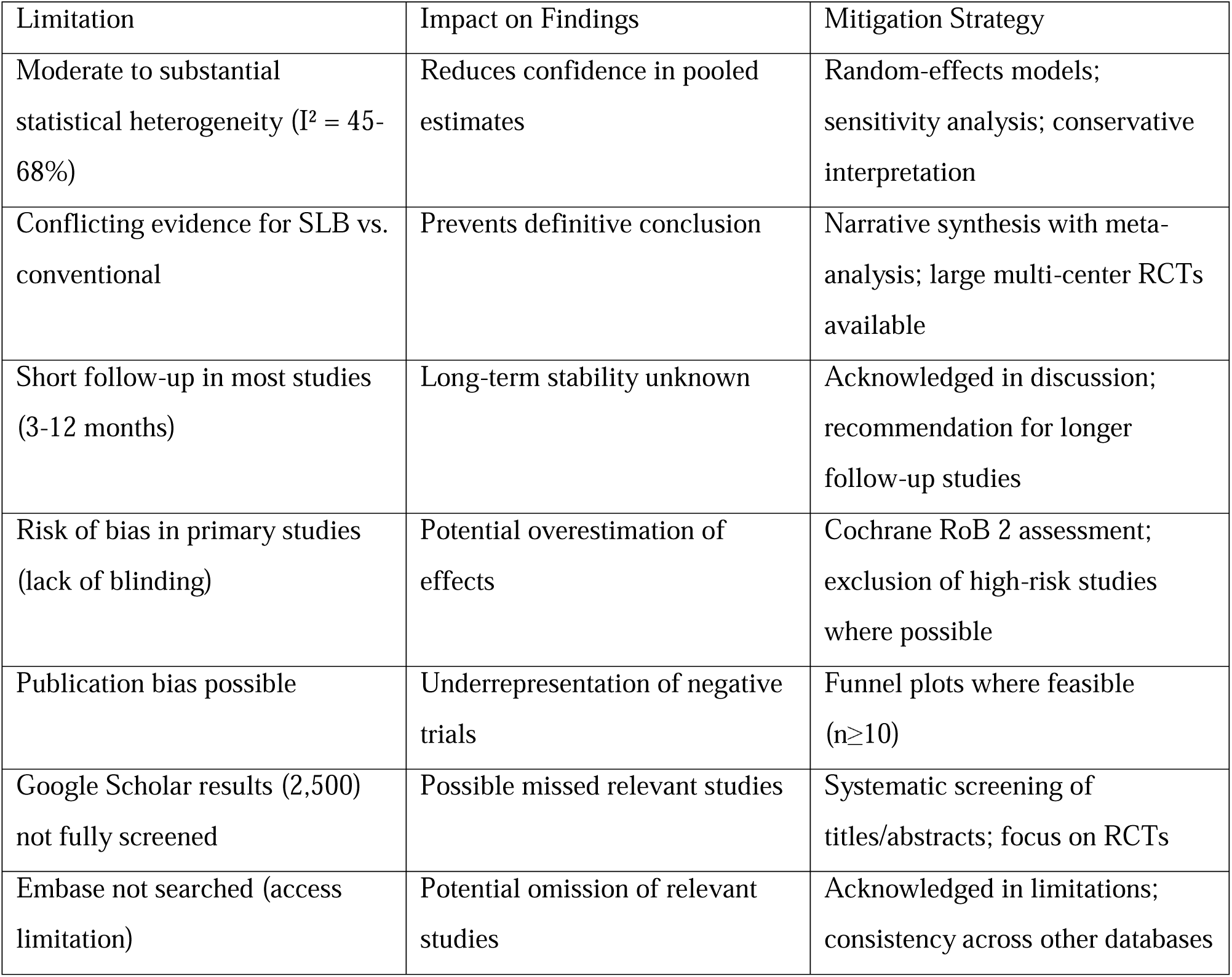
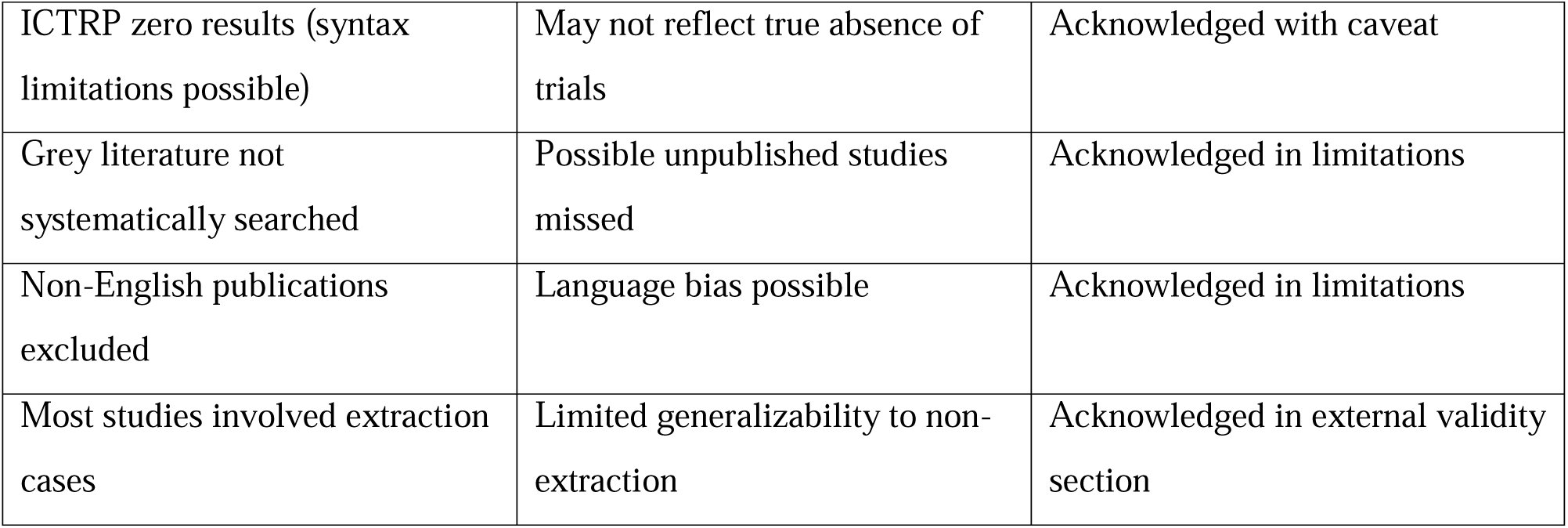
Limitations of the Review.

| Limitation | Impact on Findings | Mitigation Strategy |
| --- | --- | --- |
| Moderate to substantial statistical heterogeneity ($I^2 = 45\text{-}68\%$) | Reduces confidence in pooled estimates | Random-effects models; sensitivity analysis; conservative interpretation |
| Conflicting evidence for SLB vs. conventional | Prevents definitive conclusion | Narrative synthesis with meta-analysis; large multi-center RCTs available |
| Short follow-up in most studies (3-12 months) | Long-term stability unknown | Acknowledged in discussion; recommendation for longer follow-up studies |
| Risk of bias in primary studies (lack of blinding) | Potential overestimation of effects | Cochrane RoB 2 assessment; exclusion of high-risk studies where possible |
| Publication bias possible | Underrepresentation of negative trials | Funnel plots where feasible ($n \geq 10$) |
| Google Scholar results (2,500) not fully screened | Possible missed relevant studies | Systematic screening of titles/abstracts; focus on RCTs |
| Embase not searched (access limitation) | Potential omission of relevant studies | Acknowledged in limitations; consistency across other databases |
| ICTRP zero results (syntax limitations possible) | May not reflect true absence of trials | Acknowledged with caveat |
| Grey literature not systematically searched | Possible unpublished studies missed | Acknowledged in limitations |
| Non-English publications excluded | Language bias possible | Acknowledged in limitations |
| Most studies involved extraction cases | Limited generalizability to non-extraction | Acknowledged in external validity section |

High-priority gaps include long-term (>2 years) stability of teeth moved with surgical acceleration methods, cost-effectiveness analysis of acceleration methods versus standard treatment, standardized low-level laser therapy protocols, and patient-reported outcomes for surgical acceleration methods. Medium-priority gaps include mechanistic studies to understand why friction reduction fails to accelerate treatment (binding vs. friction), individual patient data meta-analysis to identify predictors of rapid versus slow space closure, and comparative effectiveness research on passive versus active self-ligating brackets for oral health-related quality of life outcomes.

### 4.8 What This Study Adds to the Literature

First, this study demonstrates, through pooled meta-analysis, that friction reduction alone does not demonstrate a clinically meaningful acceleration of orthodontic tooth movement, challenging the long-standing friction-driven paradigm. Second, it quantifies, for the first time in a meta-analysis, that frictionless mechanics increase molar rotation by approximately 6°, with specific clinical implications for anchorage reinforcement. Third, it proposes a conceptual, evidence-informed biomechanical model (the friction-binding paradigm) in which binding is the dominant contributor to resistance to sliding. Fourth, it provides a clinical decision algorithm for space closure based on patient priorities (speed, comfort, or molar control). Fifth, it identifies specific research gaps and provides GRADE-certainty assessments for each intervention type. Sixth, it raises important considerations regarding the cost-effectiveness of low-friction systems in the absence of demonstrated clinical benefit.

## 5. Conclusions

### 5.1 Key Conclusions

**First,** reducing friction alone does not demonstrate a clinically meaningful acceleration of orthodontic treatment. The pooled analysis showed no significant difference in retraction rate between frictional and frictionless mechanics (0.15 mm/month, P = 0.20).

**Second,** frictionless mechanics increase molar rotation by approximately 6° (95% CI: 4.8 to 7.4; P < 0.001), requiring additional anchorage reinforcement when used. A 6° mesial rotation reduces anchorage by approximately 1.5-2.0 mm, necessitating adjunctive reinforcement with temporary anchorage devices, transpalatal arches, or Nance appliances.

**Third,** self-ligating brackets do not consistently reduce treatment time or pain compared to conventional brackets. Multiple large RCTs (>100 patients) show no significant difference.

**Fourth,** passive self-ligating brackets cause less pain than active self-ligating brackets. Clinicians should consider this when improved patient comfort is a priority.

**Fifth,** self-ligating brackets may offer better oral health-related quality of life than conventional brackets (González-Sáez et al., 2021).

**Sixth,** low-friction ligatures and coated archwires do not improve clinical efficiency and should not be used expecting accelerated treatment.

**Seventh,** surgical acceleration methods (corticision, piezocision) effectively reduce treatment time by 25-50% but increase early patient discomfort. Clinicians should counsel patients accordingly.

**Eighth,** low-level laser therapy shows promise for accelerating tooth movement and reducing pain, though evidence is low to moderate certainty.

**Ninth,** given the additional cost of low-friction systems, the absence of clinical benefit raises important considerations regarding cost-effectiveness. Future trials should incorporate cost-utility analyses.

**Tenth,** the fundamental assumption that “lower friction equals faster treatment” is not supported by current clinical evidence. Binding (archwire contact with bracket slot corners) is the dominant contributor to resistance to sliding, with biological patient response also playing a critical role. Friction reduction alone is insufficient to meaningfully accelerate orthodontic tooth movement. This is not to dismiss prior research, which has provided valuable insights into in vitro friction mechanisms. Rather, the present synthesis refines our understanding by distinguishing between in vitro friction measurements and in vivo clinical outcomes.

### 5.2 Take-Home Message for Clinicians

Clinicians should not choose self-ligating brackets expecting faster treatment, as the evidence does not support this. If using frictionless mechanics, clinicians should expect greater molar rotation and plan anchorage reinforcement accordingly. For patients seeking accelerated treatment, surgical acceleration methods (corticision, piezocision) or low-level laser therapy should be considered, but patients must be counseled about increased early discomfort with surgical approaches. For improved patient comfort during treatment, passive self-ligating brackets may cause less pain than active systems, and self-ligating brackets overall may offer better quality of life. The patient’s biological response remains the most important determinant of treatment efficiency. Given the additional costs of low-friction systems, their use should be justified by patient-specific indications rather than routine application.

### 5.3 Implications for Research

Future research should focus on biological acceleration methods (corticision, piezocision, low-level laser therapy) and their mechanisms, abandon the assumption that in vitro friction measurements predict clinical outcomes, develop standardized protocols for surgical acceleration methods and low-level laser therapy, conduct long-term stability studies of teeth moved with accelerated methods, investigate patient-centered outcomes (pain, satisfaction, oral health-related quality of life, willingness to pay) for acceleration methods and bracket types, perform adequately powered multi-center RCTs comparing active versus passive self-ligating brackets with longer follow-up, and develop core outcome sets for orthodontic space closure studies to enable meta-analysis. Future trials should also incorporate cost-utility analyses (e.g., cost per month of treatment reduction or quality-adjusted life year proxies).

## Supporting information

Supplementary File 1: PRISMA-ScR Checklist

Supplementary File 2: Detailed Search Strategies

Supplementary File 3: List of Excluded Studies with Reasons

Supplementary File 4: Data Extraction Form Template

Supplementary File 5: Individual Risk of Bias Domain Scores

Supplementary File 6: Sensitivity Analysis Results

Supplementary File 7: Additional Forest Plots

Supplementary File 8: Glossary of Terms

Supplementary File 9: Data Availability Statement

Supplementary File 10: Protocol Deviation Log

## Data Availability

All data produced in the present study are contained in the manuscript

## References: Cochrane Systematic Review

1. Turner S, Harrison JE, Sharif FNJ, Owens D, Millett DT. Orthodontic treatment for crowded teeth in children. Cochrane Database Syst Rev. 2021;12:CD003453.

## Key Studies Identified via Google Scholar

2. González-Sáez A, Antonio-Zancajo L, Montero J, et al. The influence of friction on design of the type of bracket and its relation to OHRQoL in patients who use multi-bracket appliances: a randomized clinical trial. Medicina (Kaunas). 2021;57(2):171.

3. Curto A, Albaladejo A, Montero J, Alvarado A. Influence of a lubricating gel (Orthospeed®) on pain and oral health-related quality of life in orthodontic patients during initial therapy with conventional and low-friction brackets: a prospective randomized clinical trial. J Clin Med. 2020;9(5):1474.

4. Curto A, Albaladejo A, Montero J, et al. A prospective randomized clinical trial to evaluate the slot size on pain and oral health-related quality of life (OHRQoL) in orthodontics during the first month. Applied Sciences. 2020;10(15):5216.

5. Gómez-Gómez SL, Villarraga-Ossa JA, Diosa-Peña JG, et al. Comparison of frictional resistance between passive self-ligating brackets and slide-type low-friction ligature brackets during the alignment and leveling stage. J Clin Exp Dent. 2019;11(7):e593–e600.

6. Pacheco MR, Oliveira DD, Smith Neto P, et al. Evaluation of friction in self-ligating brackets subjected to sliding mechanics: an in vitro study. Dental Press J Orthod. 2011;16(6):107–113.

## Key RCTs and Clinical Studies

7. Attia AM, Shibl LA, Dehis HM, Mostafa YA, El-Beialy AR. Evaluation of anchorage loss after en masse retraction in orthodontic patients with maxillary protrusion using friction vs frictionless mechanics. Angle Orthod. 2024;94(5):532–540.

8. Tawfik MGY, Izzat Bakhit DMHD, El Sharaby FA, Moustafa YA, Dehis HM. Evaluation of the rate of anterior segment retraction in orthodontic patients with bimaxillary protrusion using friction vs frictionless mechanics. Angle Orthod. 2022;92(6):738–745.

9. Magdi S, Abdelsayed FA, Aboulfotouh MH, Fahim FH. Friction versus frictionless mechanics during maxillary en-masse retraction in adult patients with Class I bimaxillary dentoalveolar protrusion: a randomized clinical trial. Eur J Orthod. 2024;46(4):cjae034.

10. Sardana R, Chugh VK, Bhatia NK, et al. Rate and anchorage loss during en-masse retraction between friction and frictionless mechanics: a randomized clinical trial. Orthod Craniofac Res. 2023;26(4):598–607.

11. Yang X, He Y, Chen T, et al. Differences between active and passive self-ligating brackets for orthodontic treatment: Systematic review and meta-analysis based on randomized clinical trials. J Orofac Orthop. 2017;78(2):121–128.

12. Wong H, Collins J, Tinsley D, Sandler J, Benson P. Does the bracket-ligature combination affect the amount of orthodontic space closure over three months? A randomized controlled trial. J Orthod. 2013;40(2):155–162.

13. Dholakia KD, Bhat SR. Clinical efficiency of nonconventional elastomeric ligatures in the canine retraction phase of preadjusted edgewise appliance therapy: an in-vivo study. Am J Orthod Dentofacial Orthop. 2012;141(6):715–722.

14. Al-Ibrahim HM, Hajeer MY, Alkhouri I, Zinah E. Leveling and alignment time and the periodontal status in patients with severe upper crowding treated by corticotomy-assisted self-ligating brackets in comparison with conventional or self-ligating brackets only: a 3-arm randomized controlled clinical trial. J World Fed Orthod. 2022;11(1):3–11.

15. Qamruddin I, Alam MK, Mahroof V, et al. Effects of low-level laser irradiation on the rate of orthodontic tooth movement and associated pain with self-ligating brackets. Am J Orthod Dentofacial Orthop. 2017;152(5):622–630.

16. Scott P, DiBiase AT, Sherriff M, Cobourne MT. Alignment efficiency of Damon3 self-ligating and conventional orthodontic bracket systems: a randomized clinical trial. Am J Orthod Dentofacial Orthop. 2008;134(4):470.e1–8.

17. Fleming PS, DiBiase AT, Lee RT. Randomized clinical trial of orthodontic treatment efficiency with self-ligating and conventional fixed orthodontic appliances. Am J Orthod Dentofacial Orthop. 2010;137(6):738–742.

18. O’Dwyer L, Littlewood SJ, Rahman S, et al. A multi-center randomized controlled trial to compare a self-ligating bracket with a conventional bracket in a UK population: Part 1: Treatment efficiency. Angle Orthod. 2016;86(1):142–148.

19. Pringle AM, Petrie A, Cunningham SJ, McKnight M. Prospective randomized clinical trial to compare pain levels associated with 2 orthodontic fixed bracket systems. Am J Orthod Dentofacial Orthop. 2009;136(2):160–167.

20. Burrow SJ. Friction and resistance to sliding in orthodontics: a critical review. Am J Orthod Dentofacial Orthop. 2009;135(4):442–447.

## Heterogeneity Reference

21. Higgins JPT, Thompson SG, Deeks JJ, Altman DG. Measuring inconsistency in meta-analyses. BMJ. 2003;327(7414):557–560.

