## Supplementary File 1: PRISMA-ScR Checklist for "Friction in Orthodontics Revisited: A Scoping Review and Meta-Analysis Challenging the Friction-Driven Paradigm: Evidence for Binding-Dominated Resistance to Sliding"

**Authors:** Maen Mahfouz, DDS, MSc, MFD RCSI, MFDS RCSEd, MFDS RCSEng, MFDS RCPS(Glasg); Eman Alzaben, BDS

### **Supplementary File 1: PRISMA-ScR Checklist**

| Section/Item | Item Description | Reported on Page # |
| --- | --- | --- |
| **TITLE** |  |  |
| Title | Identify the report as a scoping review | 1 |
| **ABSTRACT** |  |  |
| Structured summary | Provide a structured summary including background, objectives, methods, results, conclusions | 2 |
| **INTRODUCTION** |  |  |
| Rationale | Describe the rationale for the review in the context of what is already known | 4 |
| Objectives | Provide an explicit statement of the questions and objectives being addressed | 4 |
| **METHODS** |  |  |
| Protocol | Indicate whether a review protocol exists; state if and where it can be accessed | 5 |
| Eligibility criteria | Specify characteristics of included studies | 5 |
| Information sources | Describe all information sources | 6 |
| Search | Present the full search strategy | 6 |
| Selection of sources | Describe the process for selecting sources | 7 |
| Data charting | Describe the method of data extraction | 7 |
| Data items | List and define all variables for which data were sought | 7 |
| Critical appraisal | Provide the methods used to assess risk of bias | 7 |
| Synthesis | Describe the methods for handling and summarizing data | 7-8 |
| **RESULTS** |  |  |
| Selection of sources | Give numbers of screened, assessed, and included sources | 8 |
| Characteristics of sources | Present the characteristics of included sources | 8-9 |
| Critical appraisal | Present the results of risk of bias assessment | 10 |
| Synthesis of results | Present the results of the synthesis | 10-15 |
| **DISCUSSION** |  |  |
| Summary of evidence | Summarize the main findings, including limitations | 16-22 |
| Limitations | Discuss the limitations of the review | 21-22 |
| Conclusions | Provide general interpretation of results | 22-23 |
| **FUNDING** |  |  |
| Funding | Describe sources of funding | 1 |
