## Supplementary File 2: Detailed Search Strategies for "Friction in Orthodontics Revisited: A Scoping Review and Meta-Analysis Challenging the Friction-Driven Paradigm: Evidence for Binding-Dominated Resistance to Sliding"

**Authors:** Maen Mahfouz, DDS, MSc, MFD RCSI, MFDS RCSEd, MFDS RCSEng, MFDS RCPS(Glasg); Eman Alzaben, BDS

### **Supplementary File 2: Detailed Search Strategies**

**Cochrane Library:**

#1 MeSH descriptor: [Orthodontic Brackets] explode all trees
#2 MeSH descriptor: [Orthodontic Wires] explode all trees
#3 MeSH descriptor: [Friction] explode all trees
#4 (orthodontic* NEAR/3 friction*):ti,ab,kw
#5 (sliding NEXT mechanic*):ti,ab,kw
#6 (self-ligating NEAR/3 bracket*):ti,ab,kw
#7 #1 OR #2 OR #3 OR #4 OR #5 OR #6

**PubMed:**

("Orthodontics"[Mesh] OR "Orthodontic Brackets"[Mesh]) AND
("Friction"[Mesh] OR "sliding mechanics"[Title/Abstract]) AND
("randomized controlled trial"[Publication Type] OR "clinical trial"[Publication Type])

**Google Scholar:** "orthodontic friction" OR "self-ligating brackets" AND ("RCT" OR "randomized")
