## Supplementary File 3: List of Excluded Studies with Reasons for "Friction in Orthodontics Revisited: A Scoping Review and Meta-Analysis Challenging the Friction-Driven Paradigm: Evidence for Binding-Dominated Resistance to Sliding"

**Authors:** Maen Mahfouz, DDS, MSc, MFD RCSI, MFDS RCSEd, MFDS RCSEng, MFDS RCPS(Glasg); Eman Alzaben, BDS

### **Supplementary File 3: List of Excluded Studies with Reasons**

| Study (First Author, Year) | Reason for Exclusion |
| --- | --- |
| Abdelrahman et al., 2015 | In vitro study; no clinical outcomes |
| Alavi et al., 2011 | In vitro friction measurement only |
| Alavi et al., 2012 | In vitro study |
| Alavi et al., 2013 | In vitro study |
| Alavi et al., 2015 | In vitro study |
| Akaike et al., 2015 | In vitro coating study |
| Albawardi et al., 2022 | In vitro friction measurement |
| Al-Khatib et al., 2005 | In vitro fretting study |
| Arici et al., 2021 | In vitro friction measurement |
| Bandeira et al., 2011 | In vitro coating study |
| Bandeira et al., 2020 | In vitro cytotoxicity study |
| Braun et al., 1999 | Critical review (not RCT) |
| Chang et al., 2013 | In vitro study |
| Chang et al., 2014 | In vitro study |
| da Silva et al., 2013 | In vitro coated wire study |
| da Silva et al., 2015 | In vitro study |
| de Lima Mendonça et al., 2014 | In vitro bracket protector study |
| Drescher et al., 1990 | In vitro study (German) |
| Elayyan et al., 2008 | Ex vivo study (retrieved archwires) |
| Fidalgo et al., 2011 | In vitro friction measurement |
| Gandini et al., 2013 | In vitro bracket opening/closing study |
| Ghadirian et al., 2021 | In vitro laser study |
| Gracco et al., 2019 | In vitro coating study |
| Greene et al., 2023 | In vitro friction comparison |
| Hain et al., 2003 | In vitro friction study |
| Hayashi et al., 2006 | Biomechanical modeling (not clinical) |
| Hemanth et al., 2023 | In vitro nano-coating study |
| Hodecker et al., 2022 | In vitro 3D-printed bracket study |
| Hodecker et al., 2023 | In vitro air polishing study |
| Iijima et al., 2012 | In vitro coating study |
| Kachoei et al., 2013 | In vitro nanoparticle study |
| Kawaguchi et al., 2020 | In vitro coating study |
| Khaneh Masjedi et al., 2016 | In vivo but no friction outcome (nickel release) |
| Kim et al., 2014 | In vitro friction study |
| Kopsahilis et al., 2022 | In vitro friction study |
| Krishnan et al., 2012 | In vitro coating study |
| Leite et al., 2016 | In vitro cleaning study |
| Liu et al., 2019 | Finite element analysis (not clinical) |
| Lombardo et al., 2013 | In vitro friction study |
| Major et al., 2010 | In vitro bracket tolerance study |
| Monteiro et al., 2014 | In vitro friction study |
| Moore et al., 2004 | In vitro friction study |
| Muguruma et al., 2011 | In vitro coating study |
| Muguruma et al., 2014 | In vitro torque study |
| Muguruma et al., 2017 | In vitro coating study |
| Naziris et al., 2019 | In vitro three-bracket model |
| Nucera et al., 2013 | In vitro slot design study |
| Nucera et al., 2014 | In vitro slot design study |
| Oliver et al., 2011 | In vitro friction study |
| Oz et al., 2012 | In vitro and clinical (clinical data insufficient for meta-analysis) |
| Parmagnani et al., 2012 | In vitro polishing study |
| Patil et al., 2016 | In vitro friction study |
| Pliska et al., 2011 | In vitro friction study |
| Pliska et al., 2014 | In vitro friction study |
| Reznikov et al., 2010 | In vitro friction study |
| Schlegel, 1996 | In vitro biomechanical analysis |
| Sfondrini et al., 2012 | In vitro bracket reconditioning study |
| Shahabi et al., 2017 | In vitro friction study |
| Shirakawa et al., 2018 | In vitro PEEK tube study |
| Tada et al., 2017 | In vitro PEEK wire study |
| Usui et al., 2018 | In vitro coating study |
| Voudouris et al., 2010 | In vitro friction study |
| Wichelhaus et al., 2022 | In vitro friction study |
| Williams et al., 2014 | In vitro friction study |
| Wong et al., 2014 | In vitro study |
| Yanase et al., 2014 | In vitro sliding velocity study |
| Zhang et al., 2016 | In vitro coating study |

**Total excluded from quantitative synthesis:** 1,200+ (screened out); 67 full-text articles excluded with reasons listed above.
