## Supplementary File 4: Data Extraction Form Template for "Friction in Orthodontics Revisited: A Scoping Review and Meta-Analysis Challenging the Friction-Driven Paradigm: Evidence for Binding-Dominated Resistance to Sliding"

**Authors:** Maen Mahfouz, DDS, MSc, MFD RCSI, MFDS RCSEd, MFDS RCSEng, MFDS RCPS(Glasg); Eman Alzaben, BDS

### **Supplementary File 4: Data Extraction Form Template**

#### **4.1 Study Characteristics**

| Field | Data Extracted |
| --- | --- |
| Study ID |  |
| First author |  |
| Year of publication |  |
| Country |  |
| Study design (RCT parallel / RCT split-mouth) |  |
| Sample size (total enrolled) |  |
| Sample size (analyzed) |  |
| Age range / mean (SD) |  |
| Sex distribution |  |
| Malocclusion type |  |
| Extraction pattern |  |

#### **4.2 Intervention Details**

| Field | Data Extracted |
| --- | --- |
| Intervention type |  |
| Bracket type (SLB / conventional) |  |
| SLB subtype (active / passive) |  |
| Bracket slot size (inches) |  |
| Archwire material |  |
| Archwire dimensions (inches) |  |
| Archwire cross-section (round / rectangular) |  |
| Ligation method |  |
| Force delivery system |  |
| Force magnitude (g) |  |
| Anchorage method |  |
| Acceleration method (if applicable) |  |

#### **4.3 Outcome Data**

| Outcome | Baseline (T0) | Follow-up 1 (T1) | Follow-up 2 (T2) | Follow-up 3 (T3) |
| --- | --- | --- | --- | --- |
| Space closure (mm) |  |  |  |  |
| Rate of closure (mm/month) |  |  |  |  |
| Anchorage loss (mm) |  |  |  |  |
| Molar rotation (degrees) |  |  |  |  |
| Pain (VAS, 0-10) |  |  |  |  |
| Treatment duration |  |  |  |  |

#### **4.4 Risk of Bias Assessment (Cochrane RoB 2)**

| Domain | Judgment (Low / Some / High) | Support for judgment |
| --- | --- | --- |
| Randomization process |  |  |
| Deviations from intended interventions |  |  |
| Missing outcome data |  |  |
| Measurement of outcome |  |  |
| Selection of reported result |  |  |
| Overall |  |  |
