## Supplementary File 5: Individual Risk of Bias Domain Scores for "Friction in Orthodontics Revisited: A Scoping Review and Meta-Analysis Challenging the Friction-Driven Paradigm: Evidence for Binding-Dominated Resistance to Sliding"

**Authors:** Maen Mahfouz, DDS, MSc, MFD RCSI, MFDS RCSEd, MFDS RCSEng, MFDS RCPS(Glasg); Eman Alzaben, BDS

### **Supplementary File 5: Individual Risk of Bias Domain Scores**

| Study | Randomization | Deviations | Missing Data | Outcome Measurement | Reporting | Overall |
| --- | --- | --- | --- | --- | --- | --- |
| Sardana 2023 | Low | Some | Low | Some | Low | Some concerns |
| Tawfik 2022 | Low | Low | Low | Low | Low | Low |
| Magdi 2024 | Low | Low | Low | Some | Low | Some concerns |
| Attia 2024 | Low | Low | Low | Low | Low | Low |
| Scott 2008 | Low | Low | Low | Low | Low | Low |
| Fleming 2009 | Low | Low | Low | Low | Low | Low |
| Fleming 2010 | Low | Low | Low | Low | Low | Low |
| Songra 2014 | Low | Low | Low | Low | Low | Low |
| Wahab 2012 | Low | Low | Low | Low | Low | Low |
| González-Sáez 2021 | Low | Low | Low | Low | Low | Low |
| Jung 2021 | Low | Low | Low | Low | Low | Low |
| O'Dwyer 2016 | Low | Low | Low | Low | Low | Low |
| Hoyte 2026 | Low | Low | Low | Low | Low | Low |
| Matilainen 2025 | Low | Low | Low | Low | Low | Low |
| Jahanbin 2019 | Low | Low | Low | Low | Low | Low |
| Miles 2007 | Low | Low | Low | Low | Low | Low |
| Al-Ibrahim 2022 | Low | Low | Low | Low | Low | Low |
| Qamruddin 2017 | Low | Low | Low | Low | Low | Low |
| Pringle 2009 | Low | Low | Low | Low | Low | Low |

**Notes:**

- "Some concerns" primarily due to lack of blinding in outcome assessment (unavoidable in orthodontic trials comparing bracket types)
- No studies rated as high risk of bias
- Cochrane review (CD003453) assessed as low risk of bias using AMSTAR-2
