## Supplementary File 6: Sensitivity Analysis Results for "Friction in Orthodontics Revisited: A Scoping Review and Meta-Analysis Challenging the Friction-Driven Paradigm: Evidence for Binding-Dominated Resistance to Sliding"

**Authors:** Maen Mahfouz, DDS, MSc, MFD RCSI, MFDS RCSEd, MFDS RCSEng, MFDS RCPS(Glasg); Eman Alzaben, BDS

### **Supplementary File 6: Sensitivity Analysis Results**

**6.1 Heterogeneity Classification (Non-Overlapping Thresholds)**

| I² Value | Heterogeneity Classification |
| --- | --- |
| 0-40% | Low |
| 40-60% | Moderate |
| 60-90% | Substantial |
| 90-100% | Considerable |

**6.2 Leave-One-Out Meta-Analysis for Retraction Rate**

| Excluded Study | Pooled MD (95% CI) | I² | Heterogeneity |
| --- | --- | --- | --- |
| None (primary) | 0.15 (-0.08, 0.38) | 68% | Substantial |
| Exclude Sardana 2023 | 0.18 (0.03, 0.33) | 0% | Low |
| Exclude Tawfik 2022 | 0.13 (-0.07, 0.33) | 75% | Substantial |
| Exclude Magdi 2024 | 0.15 (-0.11, 0.41) | 79% | Substantial |

**6.3 Leave-One-Out Meta-Analysis for Molar Rotation**

| Excluded Study | Pooled MD (95% CI) | I² | Heterogeneity |
| --- | --- | --- | --- |
| None (primary) | 6.10 (4.80, 7.40) | 45% | Moderate |
| Exclude Tawfik 2022 | 6.00 (4.20, 7.80) | N/A | N/A |
| Exclude Attia 2024 | 7.10 (5.30, 8.90) | N/A | N/A |

**6.4 Sensitivity Analysis Excluding High Risk of Bias Studies**

| Outcome | Excluding Some Concerns | Primary Estimate | Difference | Heterogeneity |
| --- | --- | --- | --- | --- |
| Retraction rate (MD) | 0.20 (0.05, 0.35) | 0.15 (-0.08, 0.38) | +0.05 | Substantial (68%) |
| Molar rotation (MD) | 6.10 (4.80, 7.40) | 6.10 (4.80, 7.40) | No change | Moderate (45%) |
