## Supplementary File 7: Additional Forest Plots for "Friction in Orthodontics Revisited: A Scoping Review and Meta-Analysis Challenging the Friction-Driven Paradigm: Evidence for Binding-Dominated Resistance to Sliding"

**Authors:** Maen Mahfouz, DDS, MSc, MFD RCSI, MFDS RCSEd, MFDS RCSEng, MFDS RCPS(Glasg); Eman Alzaben, BDS

### **SUPPLEMENTARY FILE 7: Additional Forest Plots**

#### **7.1 Forest Plot – Anchorage Loss: Frictional vs. Frictionless Mechanics**

**Data Table:**

| Study | Friction Mean (SD) | N | Frictionless Mean (SD) | N | MD (95% CI) | Weight (%) |
| --- | --- | --- | --- | --- | --- | --- |
| Sardana 2023 | 2.28 (0.45) | 18 | 1.13 (0.26) | 18 | -1.15 (-1.68, -0.62) | 31.8 |
| Tawfik 2022 | 1.63 (0.36) | 15 | 1.63 (0.40) | 15 | 0.00 (-0.49, 0.49) | 35.6 |
| Magdi 2024 | 1.18 (0.72) | 15 | 1.29 (0.55) | 15 | 0.11 (-0.70, 0.92) | 32.6 |

**Pooled Result (Random Effects):**

MD = -0.64 mm (95% CI: -1.32 to 0.04)
Z = 1.84 (P = 0.07)
I² = 72% (Substantial heterogeneity)

**Interpretation:** No statistically significant difference in anchorage loss between frictional and frictionless mechanics (P = 0.07). The trend favors frictionless mechanics (0.64 mm less loss), but the 95% confidence interval crosses zero. Substantial heterogeneity (I² = 72%) reflects true clinical heterogeneity across studies rather than methodological inconsistency alone. Differences in wire materials, force magnitudes, and outcome measurement methods likely contributed to this variability.

#### **7.2 Forest Plot – Root Resorption: Frictional vs. Frictionless Mechanics**

**Data Table:**

| Study | Tooth Type | Friction Mean (SD) | N | Frictionless Mean (SD) | N | MD (95% CI) |
| --- | --- | --- | --- | --- | --- | --- |
| Bakhit 2025 | Upper incisors (UI) | 0.73 (0.36) | 15 | 1.27 (0.55) | 15 | 0.54 (-0.02, 1.07) |
| Bakhit 2025 | Lower incisors (LI) | 0.70 (0.24) | 15 | 1.09 (0.55) | 15 | 0.39 (-0.11, 0.89) |

**Interpretation:** Statistically significant difference for upper incisors (MD 0.54 mm, 95% CI: -0.02 to 1.07, P = 0.06) but likely not clinically significant. For lower incisors, the difference was not statistically significant (MD 0.39 mm, 95% CI: -0.11 to 0.89, P = 0.13). No pooled I² calculated due to single study. The likelihood of root resorption should be considered when frictionless mechanics are used for retraction of incisors.

#### **7.3 Publication Bias Assessment (Pain Outcomes)**

**Number of studies:** 6

**Methodological note:** According to the Cochrane Handbook for Systematic Reviews of Interventions, funnel plots are not recommended for assessing publication bias when there are fewer than 10 studies, as the power to detect asymmetry is insufficient. Therefore, a formal funnel plot is not presented here.

**Data Summary:**

| Study | N | SMD (95% CI) | Direction |
| --- | --- | --- | --- |
| Fleming 2009 | 66 | 0.04 (-0.44, 0.52) | Neutral |
| Scott 2008 | 62 | 0.04 (-0.46, 0.54) | Neutral |
| Pringle 2009 | 52 | -0.45 (-0.85, -0.05) | Favors SLB |
| Rahman 2016 | 135 | 0.30 (-0.10, 0.70) | Favors conventional |
| Lai 2017 | 88 | -0.09 (-0.51, 0.33) | Neutral |
| González-Sáez 2021 | 90 | -0.65 (-1.10, -0.20) | Favors SLB |

**Assessment approach:** Publication bias was assessed qualitatively by examining:

1. **The direction of effect across studies:** The six studies showed mixed directions (two favoring SLB, one favoring conventional brackets, three neutral), with a pooled SMD close to zero (-0.18) and a 95% confidence interval crossing zero (-0.56 to 0.20). This diversity of results across multiple research groups and settings suggests that publication bias is unlikely to have substantially influenced the pooled estimate.
2. **Sample size range:** Studies ranged from 52 to 135 participants, with no clear relationship between sample size and effect direction.
3. **Source of studies:** Studies were identified from multiple databases (Cochrane, PubMed, Google Scholar) and included both published and registry-identified trials.

**Pooled I² for this analysis:** 68% (substantial heterogeneity)

**Conclusion:** While publication bias cannot be completely excluded, the heterogeneity of results and the absence of a clear pattern suggesting missing studies indicates that publication bias is unlikely to have substantially altered the conclusions of this meta-analysis.

#### **Summary of Heterogeneity Classifications in Supplementary File 7**

| Plot | I² | Heterogeneity Classification (Non-Overlapping Thresholds) |
| --- | --- | --- |
| 7.1 Anchorage Loss | 72% | Substantial |
| 7.2 Root Resorption | N/A (single study) | Not applicable |
| 7.3 Pain Publication Bias | 68% | Substantial |

#### **References for Supplementary File 7**

1. Sardana R, Chugh VK, Bhatia NK, et al. Rate and anchorage loss during en-masse retraction between friction and frictionless mechanics: a randomized clinical trial. Orthod Craniofac Res. 2023;26(4):598-607.
2. Tawfik MGY, Izzat Bakhit DMHD, El Sharaby FA, Moustafa YA, Dehis HM. Evaluation of the rate of anterior segment retraction in orthodontic patients with bimaxillary protrusion using friction vs frictionless mechanics. Angle Orthod. 2022;92(6):738-745.
3. Magdi S, Abdelsayed FA, Aboulfotouh MH, Fahim FH. Friction versus frictionless mechanics during maxillary en-masse retraction in adult patients with Class I bimaxillary dentoalveolar protrusion: a randomized clinical trial. Eur J Orthod. 2024;46(4):cjae034.
4. Bakhit DMI, Tawfik MGY, Dehis HM, Mostafa YA, El Sharaby FA. Position and root resorption of the incisors following anterior segment retraction using friction versus frictionless mechanics: a randomised controlled trial. J Orthod. 2025;52(1):12-21.
5. Fleming PS, Dibiase AT, Sarri G, Lee RT. Pain experience during initial alignment with a self-ligating and a conventional fixed orthodontic appliance system. A randomized controlled clinical trial. Angle Orthod. 2009;79(1):46-50.
6. Scott P, Sherriff M, Dibiase AT, Cobourne MT. Perception of discomfort during initial orthodontic tooth alignment using a self-ligating or conventional bracket system: a randomized clinical trial. Eur J Orthod. 2008;30(3):227-232.
7. Pringle AM, Petrie A, Cunningham SJ, McKnight M. Prospective randomized clinical trial to compare pain levels associated with 2 orthodontic fixed bracket systems. Am J Orthod Dentofacial Orthop. 2009;136(2):160-167.
8. Rahman S, Spencer RJ, Littlewood SJ, O'Dywer L, Barber SK, Russell JS. A multicenter randomized controlled trial to compare a self-ligating bracket with a conventional bracket in a UK population: Part 2: Pain perception. Angle Orthod. 2016;86(1):149-156.
9. Lai TT, Chiou JY, Lai TC, Chen T, Wang HY, Li CH, Chen MH. Perceived pain for orthodontic patients with conventional brackets or self-ligating brackets over 1 month period: A single-center, randomized controlled clinical trial. J Formos Med Assoc. 2020;119(1 Pt 2):282-289.
10. González-Sáez A, Antonio-Zancajo L, Montero J, et al. The influence of friction on design of the type of bracket and its relation to OHRQoL in patients who use multi-bracket appliances: a randomized clinical trial. Medicina (Kaunas). 2021;57(2):171.
11. Higgins JPT, Thompson SG, Deeks JJ, Altman DG. Measuring inconsistency in meta-analyses. BMJ. 2003;327(7414):557-560. (For funnel plot guidance)
