## Supplementary File 8: Glossary of Terms for "Friction in Orthodontics Revisited: A Scoping Review and Meta-Analysis Challenging the Friction-Driven Paradigm: Evidence for Binding-Dominated Resistance to Sliding"

**Authors:** Maen Mahfouz, DDS, MSc, MFD RCSI, MFDS RCSEd, MFDS RCSEng, MFDS RCPS(Glasg); Eman Alzaben, BDS

### **Supplementary File 8: Glossary of Terms**

| Term | Definition |
| --- | --- |
| **Binding** | Archwire contact with bracket slot corners that occurs when tooth angulation exceeds the critical contact angle; the dominant component of resistance to sliding |
| **Friction** | Surface resistance that opposes relative motion between the archwire and bracket slot |
| **Notching** | Permanent deformation of the archwire due to force overload; rare |
| **Self-ligating bracket (SLB)** | Bracket with an integrated clip or slide mechanism that secures the archwire without elastomeric or steel ligatures |
| **Active SLB** | Self-ligating bracket with a clip that applies active force to the archwire |
| **Passive SLB** | Self-ligating bracket with a clip that holds the archwire without applying active force |
| **Sliding mechanics** | Orthodontic space closure technique where the archwire slides through bracket slots |
| **Frictionless mechanics** | Space closure technique using loops (e.g., T-loops, mushroom loops) that eliminate sliding |
| **En masse retraction** | Simultaneous retraction of all anterior teeth as a single segment |
| **Anchorage loss** | Unwanted mesial movement of posterior teeth during anterior retraction |
| **Molar rotation** | Rotation of the first molar around its long axis during space closure |
| **Temporary anchorage device (TAD)** | Mini-screw implant used as skeletal anchorage |
| **Corticision** | Minimally invasive surgical procedure involving cortical bone cuts to accelerate tooth movement |
| **Piezocision** | Surgical technique using piezoelectric instruments to create cortical bone cuts |
| **Micro-osteoperforation (MOP)** | Small perforations in cortical bone to accelerate tooth movement |
| **Low-level laser therapy (LLLT)** | Photobiomodulation using low-power lasers to accelerate tooth movement and reduce pain |
| **OHRQoL** | Oral health-related quality of life |
| **VAS** | Visual Analog Scale (for pain measurement) |
| **I²** | Statistical measure of heterogeneity in meta-analysis (0-100%) |
