## Supplementary File 9: Data Availability Statement for "Friction in Orthodontics Revisited: A Scoping Review and Meta-Analysis Challenging the Friction-Driven Paradigm: Evidence for Binding-Dominated Resistance to Sliding"

**Authors:** Maen Mahfouz, DDS, MSc, MFD RCSI, MFDS RCSEd, MFDS RCSEng, MFDS RCPS(Glasg); Eman Alzaben, BDS

### **Supplementary File 9: Data Availability Statement for Supplementary Material**

All supplementary material produced in the present study is available upon reasonable request to the corresponding author. The search strategies, data extraction forms, and risk of bias assessments are provided in this supplementary file. Raw data extracted from included studies are available in the main manuscript tables.

**Contact:**
