## Supplementary File 10: Protocol Deviation Log for "Friction in Orthodontics Revisited: A Scoping Review and Meta-Analysis Challenging the Friction-Driven Paradigm: Evidence for Binding-Dominated Resistance to Sliding"

**Authors:** Maen Mahfouz, DDS, MSc, MFD RCSI, MFDS RCSEd, MFDS RCSEng, MFDS RCPS(Glasg); Eman Alzaben, BDS

### **Supplementary File 10: Protocol Deviation Log**

| # | Deviation | Description | Justification |
| --- | --- | --- | --- |
| 1 | Protocol not prospectively registered | Acknowledged as limitation | Protocol was predefined prior to data extraction |
| 2 | Embase not searched | Access limitation | Acknowledged in limitations; consistency across other databases |
| 3 | Google Scholar results screened not fully extracted | Pragmatic decision due to volume | Systematic screening applied; focus on RCTs |
| 4 | Non-English publications excluded | Language limitation | Acknowledged in limitations |
| 5 | Heterogeneity anticipated | Random-effects model used | Appropriate statistical handling of expected clinical heterogeneity |
